# Effects of bariatric surgery on functional connectivity of the reward and default mode network: a pre-registered analysis

**DOI:** 10.1101/2021.04.01.21254543

**Authors:** Hannah S. Heinrichs, Frauke Beyer, Evelyn Medawar, Kristin Prehn, Jürgen Ordemann, Agnes Flöel, A. Veronica Witte

## Abstract

Obesity imposes serious health risks and involves alterations in resting-state functional connectivity of brain networks involved in eating behavior. Bariatric surgery is an effective treatment, but its effects on functional connectivity are still under debate. In this pre-registered study, we aimed to determine the effects of bariatric surgery on major resting-state brain networks (reward and default mode network) in a longitudinal controlled design. 33 bariatric surgery patients and 15 obese waiting-list control patients (37 females; aged 44.15 ± 11.86 SD years (range 21-68)) underwent magnetic resonance imaging at baseline, after 6 and 12 months. We conducted a pre-registered whole-brain time-by-group interaction analysis, and a time-by-group interaction analysis on within-network connectivity (https://osf.io/f8tpn/, https://osf.io/59bh7/). In exploratory analyses, we investigated the effects of weight loss and head motion. Bariatric surgery compared to waiting did not significantly affect functional connectivity (FWE-corrected *p >* 0.05), neither whole-brain nor within-network. In exploratory analyses, surgery-related BMI decrease (FWE-corrected *p* = 0.041) and higher average head motion (FWE-corrected *p* = 0.021) resulted in significantly stronger connectivity of the reward network with medial posterior frontal regions. This pre-registered well-controlled study did not support a strong effect of bariatric surgery, compared to waiting, on major resting-state brain networks after 6 months. Exploratory analyses indicated that head motion might have confounded the effects. Data pooling and more rigorous control of within-scanner head motion during data acquisition are needed to substantiate effects of bariatric surgery on brain organization.

## Introduction

Obesity is a worldwide health issue, entailing huge personal and societal costs. Excess amount of body fat not only affects cardiovascular and metabolic health, but also increases the risk for cognitive decline and dementia later in life (Albanese et al., 2017). Conservative treatment options including behavioral therapy often do not yield the desired weight loss, especially in patients with very high BMI (> 35 kg/m^2^). Here, bariatric surgery is a viable option to rapidly induce weight loss and improve glycemic status. RYBG is the most invasive, yet most effective surgical procedure, but other techniques like vertical sleeve gastrectomy (VSG) and gastric banding (GB) are also common (Berthoud et al., 2011).

Bariatric surgery leads to weight loss due to malabsorption of nutrients, reduced digestion efficiency and altered food perception, appetite and central regulation of food intake (Brutman et al., 2019; Mulla et al., 2017). Precise mechanisms how bariatric surgery leads to altered appetitive signaling are yet to be elucidated. One option to address these questions on brain-behavior relationships is to use resting-state functional magnetic resonance imaging (rsfMRI), a technique capturing the dynamic organization of the brain. Functional connectivity (FC) networks, i.e. brain regions with correlated neural activity over time, are in anatomical correspondence with specific brain networks involved in cognitive processes, including attention and executive control (Smith et al., 2009). The reward network (RN), processing hedonic value and internal motivation, and the default mode network (DMN), a higher-order network, involved in interoception and governing shifts between external-internal processes, are promising candidates to mediate altered central regulation of food intake after bariatric surgery.

The RN comprises the ventromedial prefrontal cortex (vmPFC), the nucleus accumbens (NAcc), the putamen, the amygdala and the anterior insula (Liu et al., 2011; O’Doherty, 2004). These brain regions have been suggested to guide food valuation processes and decision-making in humans (Bartra et al., 2013; Hare et al., 2011; Hutcherson et al., 2012; Schmidt et al., 2018). Frequently, obesity has been associated with hyperactivation of RN regions during anticipation of (high-caloric) food cues, and in contrast, reduced activation to actual taste of these foods (Devoto et al., 2018; García-García et al., 2014; Meng et al., 2020; Stoeckel et al., 2009, though see Morys et al., 2020). RsfMRI studies also showed increased local FC of reward network regions, i.e. NAcc, vmPFC, putamen, insula (Contreras-Rodríguez et al., 2017; Coveleskie et al., 2015; Hogenkamp et al., 2016), and altered connectivity with salience, homeostatic and sensorimotor networks (Lips et al., 2014; Wijngaarden et al., 2015). In bariatric surgery patients, connectivity within the RN (e.g. putamen, OFC) might be normalized by the surgery, however the evidence is limited due to a lack of longitudinal obese control groups (Duan et al., 2020; Schmidt et al., 2021; Wiemerslage et al., 2016). Possibly, a reconfiguration of the fronto-striatal brain networks could emerge from altered gut signaling, e.g. changes in ghrelin levels, via hypothalamic-striatal projections (Karra et al., 2013; Li et al., 2019, but Zoon et al., 2018).

The DMN includes the posterior cingulate cortex (PCC)/precuneus, the medial prefrontal cortex (mPFC) and the inferior and lateral parietal cortex (Raichle, 2015). Higher BMI and obesity have been associated with a pattern of decreased within DMN FC and increased FC of DMN regions to other networks, i.e. salience and sensory networks in several resting-state and task-based rsfMRI studies (Beyer et al., 2017; Borowitz et al., 2020; Chao et al., 2018; Ding et al., 2020; Doucet et al., 2017; Kullmann et al., 2011; Sadler et al., 2018; Wijngaarden et al., 2015). After bariatric surgery, a normalization of the connectivity between DMN and cognitive control and salience brain regions might occur, yet no study has included a longitudinal control group (Frank et al., 2013; Li et al., 2018; Olivo et al., 2017). In sum, while there is some evidence hinting to a role for DMN and RN FC in altered regulation of food intake after bariatric surgery, the existing evidence is inconclusive. Most studies have investigated small cohorts of patients, without adequate obese control groups, and did not rigorously separate confirmatory from exploratory analyses (George et al., 2016).

Further, while higher BMI has been consistently associated with more head motion during rsfMRI (Beyer et al., 2017; Hodgson et al., 2016), previous studies in bariatric surgery patients have not taken this important confounder of FC into account. In the present sample, we previously reported a group-by-time interaction on head motion (Beyer et al., 2020). Thus, we aimed to rigorously control for motion-related variance in our analyses. We had the following confirmatory hypotheses:

1. Whole-brain FC of the RN and DMN changes differently from baseline to followup in the bariatric surgery compared to a waiting-list control group.
2. Within-network FC of the RN and DMN changes differently from baseline to followup in the bariatric surgery compared to a waiting-list control group.

We tested hypothesis 1 by investigating the interaction of bariatric surgery and time on DMN and RN whole-brain FC. We pre-registered two denoising pipelines, and three covariate schemes. For hypothesis 2, we performed a confirmatory analysis of the group-by-time interaction on aggregated, within-network FC, for two time points and the same covariate schemes. In exploratory analyses, we examined the whole-brain interaction effect for three time points, the effects of head motion on FC and whether weight loss, a proxy of treatment success, predicted changes in FC.

## Methods

### Sample and Study Design

The ADIPOSITAS-Study investigated the effects of bariatric surgery on brain structure and function in a prospective design at the Charité University Medicine Berlin, Germany. For more details see (Prehn et al., 2020). We used all data acquired until April 2019. The study protocol was in accordance with the Helsinki Declaration and approved by the ethics committee of the Charité University Medicine Berlin. The study was registered at clinicaltrials.gov as NCT01554228. We pre-registered the present resting-state analyses on the OSF (https://osf.io/yp42s). We made additional changes (see https://osf.io/59bh7/) to the pre-registration after preprocessing the rsfMRI data as we realized some aspects of the analysis were inadequately described in the initial pre-registration. For a comparison of the pre-registration and the manuscript, please visit https://osf.io/45n9f/. Participants were recruited from the Center for Bariatric and Metabolic Surgery at the Charité University Medicine Berlin. Inclusion criteria were, in accordance with German guidelines for bariatric surgery, a failure of conservative obesity treatment and either (1) a BMI > 40 km/m^2^ or (2) a BMI > 35 kg/m^2^ and at least one typical co-morbidity (e.g. type-2 diabetes, hypertension, non-alcoholic fatty liver disease)(Mechanick et al., 2013). Participants were aged between 18 and 70 years and had no history of cancer, chronic inflammatory disease and addiction, other severe untreated diseases, brain pathologies identified in the MRI scan or cognitive impairments (defined as MMSE score *<* 34). In total, 51 participants out of the originally enrolled 69 subjects received MRI. Five data points of three subjects had to be excluded due to bad anatomical image quality (see below), which led to a final data set with 101 rsfMRI sessions. The final sample entailed 48 morbidly obese individuals (37 females; aged 44.2 *±* 11.9 SD years (range 21-68)). 60.4% of participants had clinically diagnosed hypertension, 4.2% had type-2 or type-1 diabetes, and 6.2% reported to smoke.

Participants either underwent surgery (n = 33, 26 females) or were waiting list controls (n = 15, 11 females), who waited for their health insurance’s approval to undergo surgery. Measures were taken at baseline (BL), 6 (FU1) and 12 (FU2) months post-surgery/baseline appointment to capture both phases of rapid weight-loss and maintenance (Maciejewski et al., 2016). Analyses were performed on all participants who provided at least one data point of rsfMRI data. 19 participants had complete data, 15 provided data for two time points and 14 for one time point (for more details see Figure 1 in supplementary information (SI)). The pre-registered analysis of changes from baseline to followup included 24 participants with both time points, in total 72 data points.

**Figure 1.**
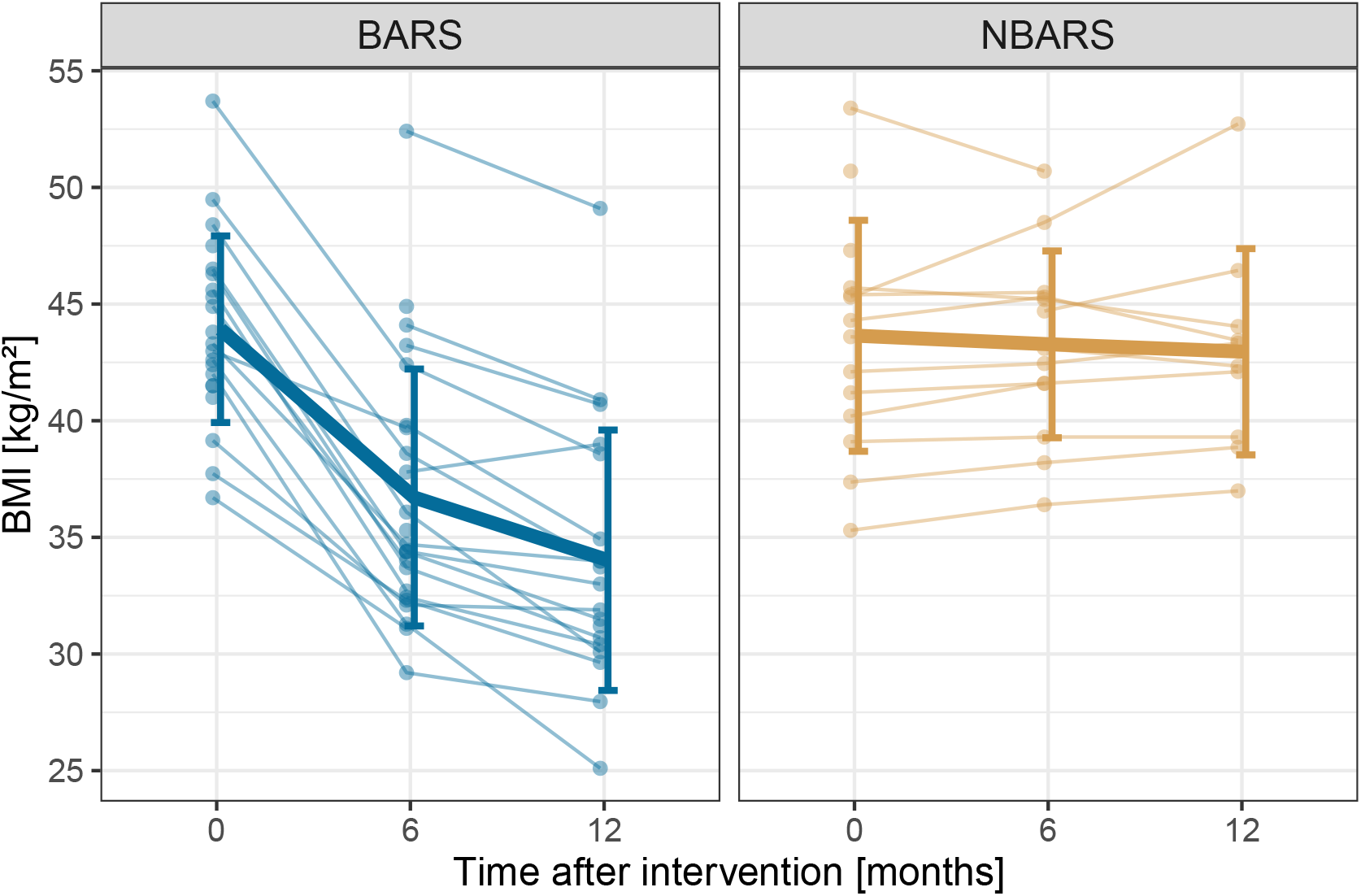
Trajectory of BMI separately for the bariatric surgery group (BARS) and the waiting-list control group (NBARS); individual trajectories are plotted in transparent, mean trajectories including standard deviations in opaque colors.

15 patients underwent RYGB, 12 underwent VSG and 1 GB, for 5 patients in the intervention group information was not available. Participants arrived in the morning (between 07:00 and 12:00 a.m.) after an overnight fast. They underwent medical assessments including an interview, blood draw and anthropometric measurements before having a one hour break for breakfast. MRI scanning was done after performing a psychological test battery (for details see Prehn et al., 2020).

### MRI Acquisition

MRI was performed with a 12-channel head coil on a 3 Tesla Trio, Siemens (Erlangen) with syngo B17 software. T1-weighted anatomical images were acquired as described in Prehn et al., 2020 (with MPRAGE, repetition time (TR) = 1900 ms, echo time (TE) = 2.52 ms, flip angle = 9°, voxel size= 1 *×* 1 *×* 1 mm^3^, 192 sagital slices). Resting-state echo-planar imaging was acquired with a TR of 2.3 s and TE of 30 ms. The image matrix was 64 *×* 64 with an in-plane resolution of 3 mm *×* 3 mm and 34 slices with a slice thickness of 4 mm. 150 volumes were acquired, resulting in a total acquisition time of 5:45 minutes. Additionally, a gradient echo field map with a TE difference of 2.46 ms was acquired to correct for field inhomogeneities. Participants were instructed to close their eyes but to remain awake during scanning.

### Preprocessing

### Minimal Preprocessing

Imaging data analysis was conducted using AFNI 19.1.05, ANTS 2.3.1, FSL 6.0.1 and FreeSurfer 6.0.0p1, wrapped in a nipype workflow (version 1.2.0) in Python 2.7.15 which can be found on https://github.com/fBeyer89/ADI_preproc/. T1-weighted images were first processed by FreeSurfer’s cross-sectional pipeline (Fischl, 2012). Then, Freesurfer’s longitudinal stream was applied to all cross-sectional runs (Reuter & Fischl, 2011). Here, white matter (WM) and cerebral spinal fluid (CSF) masks were derived based on FreeSurfer’s `aseg.mgz` segmentation file for quality control of rsfMRI preprocessing. The skull-stripped brain (`brainmask.mgz`) was then coregistered to the MNI152 1 *×* 1 *×* 1 mm template using ANTS (Avants et al., 2009). Minimal functional preprocessing included the removal of first four volumes, motion correction (FSL’s MCFLIRT), fieldmap distortion correction (FSL’s `fsl_prepare_fieldmap` and `FUGUE`) and coregistration to the subject’s individual longitudinal anatomical space (FreeSurfer’s `bbregister`). In more detail, the transformations derived from the latter three steps were combined into one and applied in a single step. For further analysis and ICA-AROMA processing, the minimally preprocessed data were intensity normalized and smoothed with a 6 mm Gaussian kernel (`fslmaths-kernel gauss 2.548`).

### Denoising Pipelines

In the pre-registration, we specified two denoising pipelines (ICA-AROMA and CompCor (AROMA+CC) and ICA-AROMA, CompCor and global signal regression (AROMA+CC+GSR)), for details see SI (Ciric et al., 2017; Parkes et al., 2018).

### Quality Assessment

The quality of anatomical images and rsfMRI was assessed separately. To control the quality of the anatomical imaged, FreeSurfer cross-sectional and longitudinal segmentations were visually checked according to Klapwijk et al., 2019. We excluded 5 datasets from 3 participants because of excessive head motion leading to failed pial reconstruction and anatomical-functional coregistration. RsfMRI quality control was performed according to the protocol by Ciric et al., 2018 (see SI for more details). Head motion was quantified using mean framewise displacement (mFD) according to Power et al., 2012 and log-transformed for further analysis (logmFD). As pre-registered, we did not exclude anybody based on high average head motion (Beyer et al., 2020).

## Functional Connectivity

### Whole Brain Functional Connectivity

To derive RN and DMN FC maps, we used NAcc and PCC/precuneus as seed regions of interest (ROI), respectively. We did not select vmPFC for the RN due to low SNR. Based on FreeSurfer’s aseg.mgz and Desikan-Killiany parcellation, we created seed masks using `mri_binarize` (thresholded for NAcc at 26,58; precuneus at 1025,2025) and averaged them over hemispheres. Then, we used `NiftiLabelsMasker` and `NiftiMasker` to extract the standardized timeseries from the seed regions and the whole brain. We calculated the Pearson’s correlation between them with numpy.dot, performed r-to-z Fisher-transformation and saved the resulting correlation maps for each preprocessing pipeline (minimally preprocessed, AROMA, AROMA+CC, AROMA+CC+GSR). Finally, the connectivity maps were transformed into MNI space using the affine transformation and non-linear warp derived with ANTS during anatomical preprocessing.

### Aggregated within-network Functional Connectivity

To extract aggregated within-network FC, we first calculated the mean DMN and RN over all participants and time points, adjusted for age and sex. We used GSR-denoised data as input and clusterwise bootstrapping with *N* = 1000. Network masks were formed from all voxels within clusters which survived a clusterwise multiple comparison correction of FWE-corrected *p* < 0.05. We extracted the average GSR-denoised FC from these masks.

## Statistical Analysis

Statistical analyses were performed in MATLAB version 9.7.0.1190202 (R2019b, MATLAB, 2018) using the SwE toolbox version 2.2.2 (Guillaume et al., 2014) as implemented in the Statistical Parametric Mapping software (SPM12.7770, Ashburner et al., 2014). The marginal model implemented in the SwE toolbox implicitly accounts for random effects without the need to specify them through the error term. We used a modified SwE assuming different covariance structures for the intervention and the control group because of their unbalanced sample size. We used an explicit brain mask, derived from the MNI ICBM “152 nonlinear 6th generation” atlas (re-sampled to 3 *×* 3 *×* 3 mm^3^ and thresholded at 0.5 GM probability) for all analyses. Statistical analyses on the aggregated FC and imputation of missing data were performed in R version 3.6.1 (Team, 2013).

### Confirmatory Analysis (pre-registered)

We tested the pre-registered hypothesis of a time-by-group interaction for two time points.

### Whole-brain analysis

As pre-registered, we performed the analysis for baseline and followup time points only, for AROMA+CC and AROMA+CC+GSR denoising pipelines and adjusting for no confounders (model CA1), age, sex and average logmFD of both time points (model CA2) and age, sex, average logmFD and baseline BMI (model CA3). Because the information about the BMI at baseline of one participant in the intervention group was missing, we employed multivariate imputation, for details see SI.

### Aggregated FC analysis

We analyzed the aggregate FC using linear mixed models in the R package lme4 (Bates et al., 2007). We deviated from the pre-registration by only investigating data from the AROMA+CC+GSR denoising pipeline. First, we investigated the time-by-group interaction for baseline and followup time points only. We either adjusted for no confounders (model CA1), for baseline age, sex, average of logmFD of both time points (model CA2) or additionally for baseline BMI (model CA3). We performed model comparison between R1 and R0 models, where `R1 = lmer(aggFC ∼ timepoint*group + age + sex + (1|subj)), and R0 = lmer(aggFC ∼ timepoint + group + age + sex + (1|subj)).` As specified in the pre-registration, we repeated the above-mentioned interaction analysis for all three time points.

## Exploratory Analysis

### Whole-brain analysis

As described in the pre-registration, we calculated the between- and within subject centered values of BMI (Guillaume et al., 2014). This model, containing average BMI and BMI change, allowed us to disentangle the differential effects of these variables on FC. We first estimated their effects in a model adjusting for baseline age and sex (model EA1) and then additionally controlling for logmFD (model EA2). As we previously reported correlated change in BMI and head motion in this sample (Beyer et al., 2019), we explored a refined model including average BMI and logmFD and change in both measures, along with baseline age and sex (model EA3). Here, we aimed to see whether any effect of change in BMI would be detectable when adjusting for the change in head motion. In addition to these pre-registered exploratory analyses, we explored our whole-brain group-by-time point interaction models for the data of all three time points on whole-brain level and for aggregated values. In this model, time was represented as factor taking into account possible non-linear time courses in the increase and decrease of FC over the course of one year, which may occur depending on the phase of weight management (Olivo et al., 2017). The resulting factorial design contained one regressor for each time point per group (see SI for depiction of design matrix). This analysis had not been pre-registered. Here, we used individual logmFD values (not averaged) as covariate to capture variance in logmFD change over time points. We investigated two models adjusting for age and sex (model EA4) and age, sex and logmFD (model EA5). For a better understanding of the unique contribution of average and longitudinal change in logmFD measures, we tested the association of head motion and FC in the additional exploratory model EA6: `FC = between-subject logmFD + within-subject logmFD` with age and sex as nuisance covariates.

### Aggregated FC analysis

Further, we performed the pre-registered exploratory analysis with average BMI and change in BMI as predictors of the aggregated FC from AROMA+CC+GSR denoised data of both networks. We calculated three models with average and change in BMI as predictors of interest and adjusted for baseline age and sex (model EA1), logmFD (model EA2) and average and change in logmFD (model EA3).

## Statistical Inference

### Whole-brain analysis

To ensure robustness of our results, we used non-parametric inference testing based on wild bootstrap with an unrestricted SwE on all contrasts of interest for clusterwise inference. Deviating from the pre-registration, we used type C2 instead of type 3 for small sample bias adjustment as this was recommended for wild bootstraps in the SwE manual. Deviating from the pre-registration but prior to the analysis, we fixed a cluster forming threshold of *p* < 0.001 for more rigorous multiple comparison adjustment (instead of *p* < 0.01), and 1000 bootstraps due to required computation time (instead of 5000). Significant clusters are defined as family-wise error (FWE) corrected *p* < 0.05. The anatomical localization of significant clusters was investigated with the SPM Anatomy toolbox, version 2.2c (Eickhoff et al., 2005) and the Harvard-Oxford Atlas in FSL version 5.0.11.

### Aggregated FC analysis

The interaction effect of group and time point in the models of aggregated FC was considered significant if the model comparison between R1 and R0 models using the anova command showed *p* < 0.05. In all exploratory models, we considered all coefficients with *p* < 0.05 as significant.

### Functional Decoding

In an exploratory analysis, we compared the resulting contrast maps with whole-brain activation maps from the NeuroSynth (https://www.neurosynth.org/) database (Yarkoni et al., 2011). We uploaded the contrast images for change BMI and average logmFD on NeuroVault, and applied the decoding classifier. This classifier estimates the similarity of meta-analytic activation maps of +500 search terms with our our contrast maps. We reported the three top terms for both contrasts.

## Results

Histograms on baseline characteristics revealed that patients in the control group did not differ notably from the intervention group regarding BMI and mFD. There were slight differences in the distribution of sex and age, the control group had a higher number of male participants (n = 4 vs. n = 10) and a higher mean age (47.14 vs. 39.33). Change in BMI throughout the study is depicted in Figure 1.

## Confirmatory analysis (pre-registered)

### Whole-brain Analysis

Against our initial hypothesis, there was no interaction effect of group and time point on neither RN nor DMN FC in model CA 1 (no adjustments). There also was no significant main effect for any of the effects of interest (time, group) in clusterwise inference with FWE-correction. The same was true for models CA2 (adjusting for age, sex and average of logmFD). Results did not differ between AROMA+CC and AROMA+CC+GSR denoising pipelines. In model CA3 (adjusting for age, sex, average of logmFD and baseline BMI) there also was no significant interaction when adding baseline BMI. Yet, we found a significant main effect of time in this model. For AROMA+CC+GSR denoised data, there was decreased FC of the NAcc with the lateral occipital cortex at baseline compared to followup (FWE-corrected *p* = 0.030), and decreased FC of the PCC within the DMN to the medial anterior cingulate cortex (FWE-corrected *p* = 0.046) (see SI). The unthresholded contrast maps of group, time and group-by-time interaction for the unadjusted model for AROMA+CC+GSR were uploaded on NeuroVault.

### Aggregated FC Analysis

There was no significant group-by-time interaction for aggregated DMN and RN FC (see Figure 2 and SI for a detailed summary of the models), regardless of the adjustments (CA1 without adjustment, CA2 adjusting for age, sex and average of mFD or CA3 age, sex, average of mFD and baseline BMI), and whether the analyses were performed in two and three time points.

**Figure 2.**
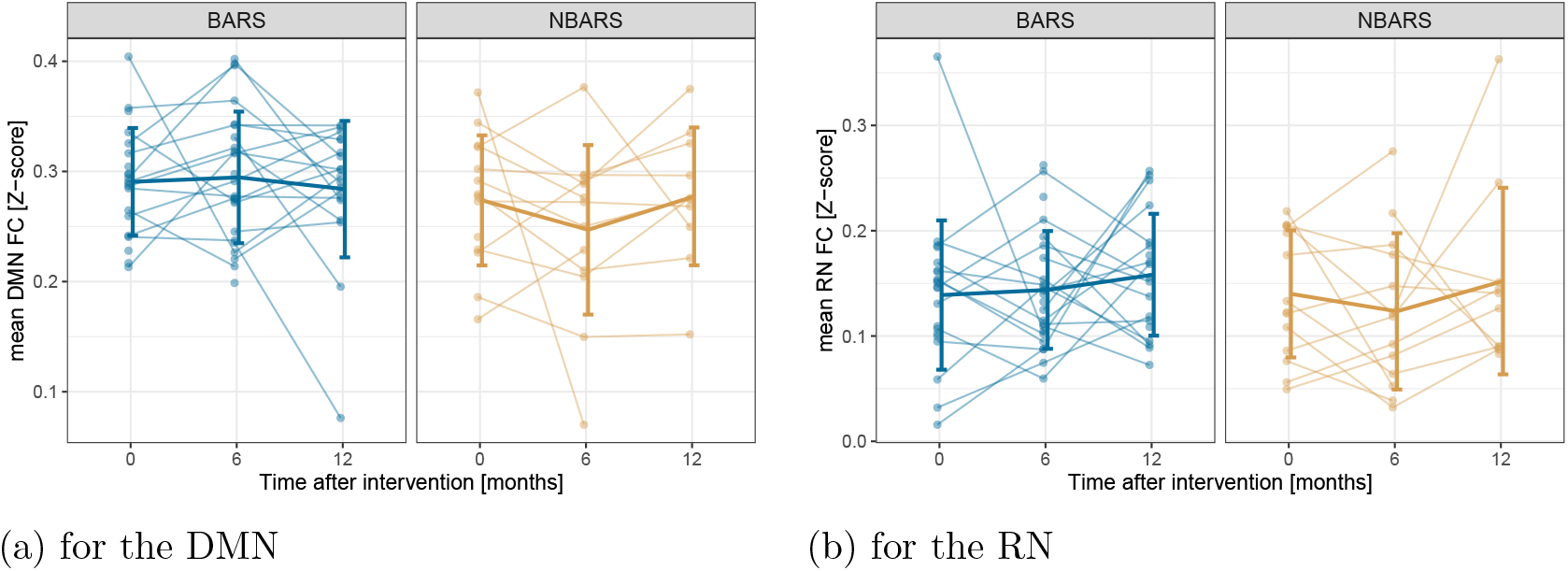
Mean network connectivity per group over time separately for the bariatric surgery group (BARS) and the waiting-list control group (NBARS); individual trajectories are plotted in transparent, mean trajectories including standard deviations in opaque colors.

## Exploratory Analysis

### Whole-brain Analysis

RN FC was not significantly related to neither average nor change in BMI, for either of the denoising pipelines and regardless whether we adjusted for logmFD in models EA1 and EA2. Only in model EA3 (adjusting for average and change in both BMI and logmFD), we found that more BMI decrease (e.g. weight loss) predicted higher FC between NAcc and a cluster in the posterior-medial frontal region (see Figure 3 and Table 2).

**Table 2.**
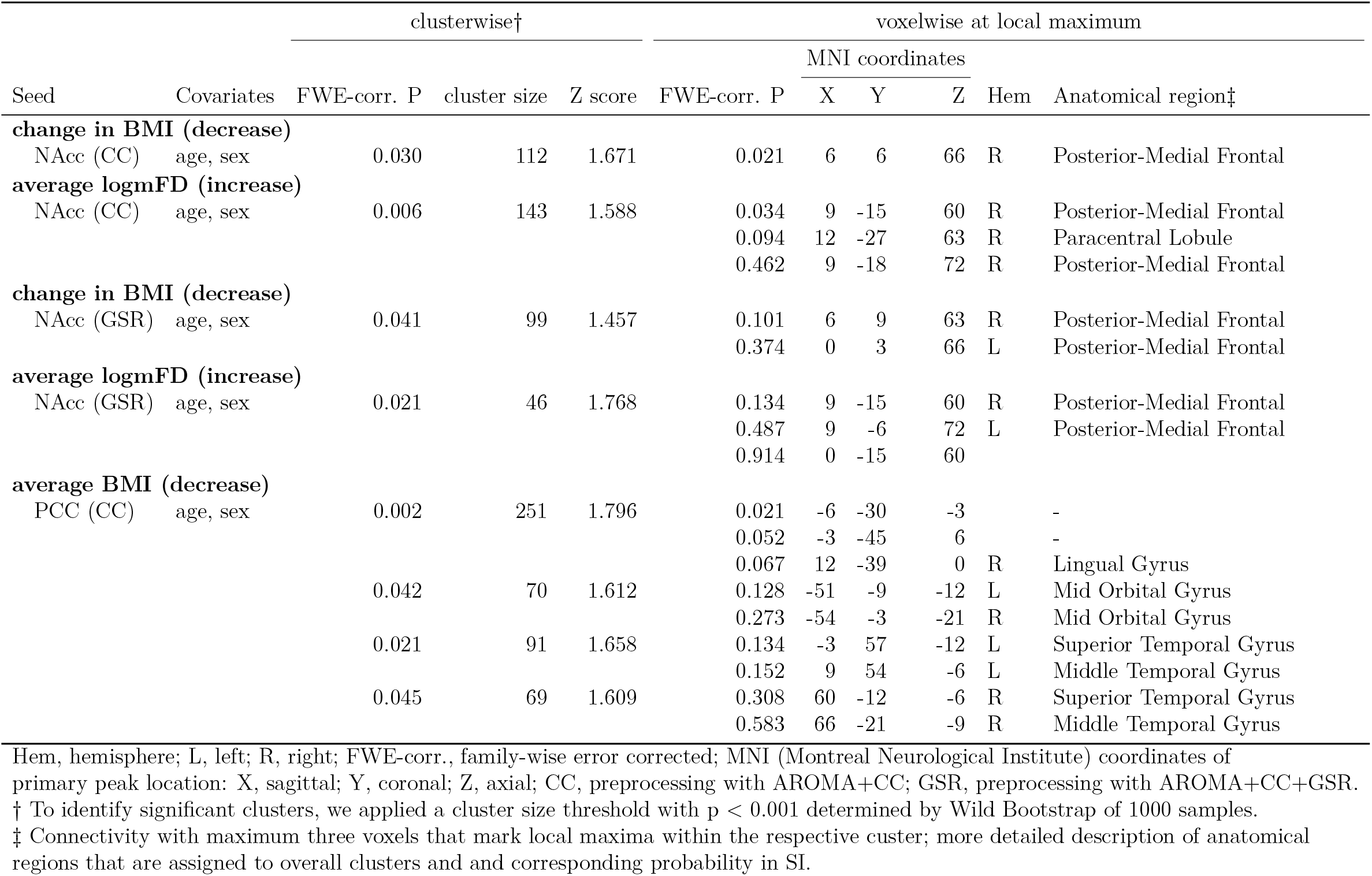
Changes in functional connectivity in whole brain analysis in model EA3

**Figure 3.**
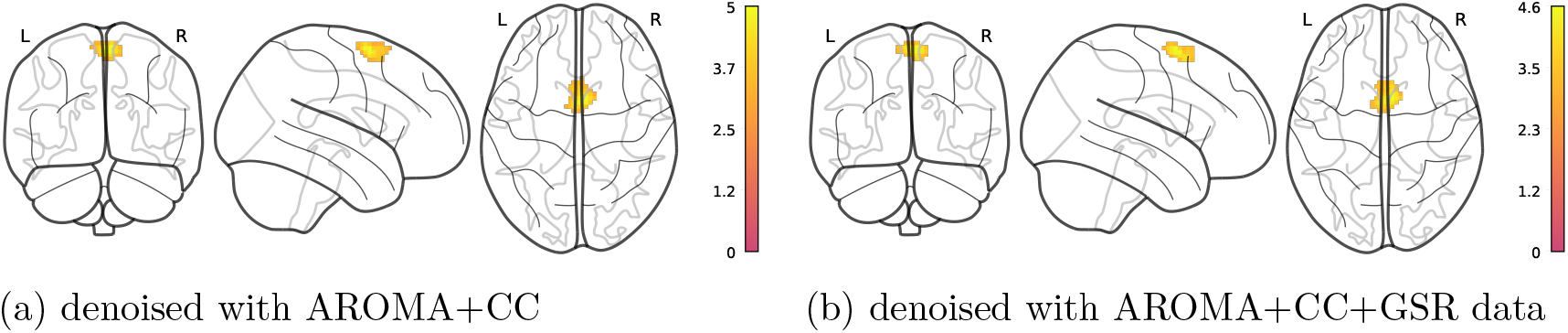
Stronger BMI decrease is associated with increased FC between NAcc and posterior-medial frontal region, adjusted for age, sex, average BMI and logmFD (model EA3). Legends denote empirical Z-values.

The peak voxel was classified as belonging to superior frontal gyrus (45% probability) and supplementary motor area (SMA) (37% probability) in the Harvard-Oxford atlas. Voxel activation at local maximum within this cluster was significant at peak level after FWE-correction (*p* = 0.030), similarly in AROMA+CC+GSR-denoised data (*p* = 0.041).

Moreover, average logmFD was positively associated with FC between NAcc and motor cortex in model EA3 for both denoising pipelines (see Figure 4 and Table 2). For the DMN, we found that higher average BMI predicted lower FC of the precuneus/PCC with the lingual gyrus, mid orbital gyrus and temporal gyrus in the images denoised with AROMA+CC. This finding was significant in models EA1, EA2 and EA3 (see Figures 5 and 6 and Table 1). Yet, none of the clusters survived when using AROMA+CC+GSR denoised data (see Table 2). Unthresholded maps for the t-tests as well as contrasts of average BMI and change BMI of the main model, and post-hoc contrasts for average logmFD and change in logmFD were published on NeuroVault. Similarly to the analysis with two time points, there was no significant interaction or main effect when analyzing models EA4 and EA5 in the full data set of three time points for neither RN nor DMN.

**Table 1.**
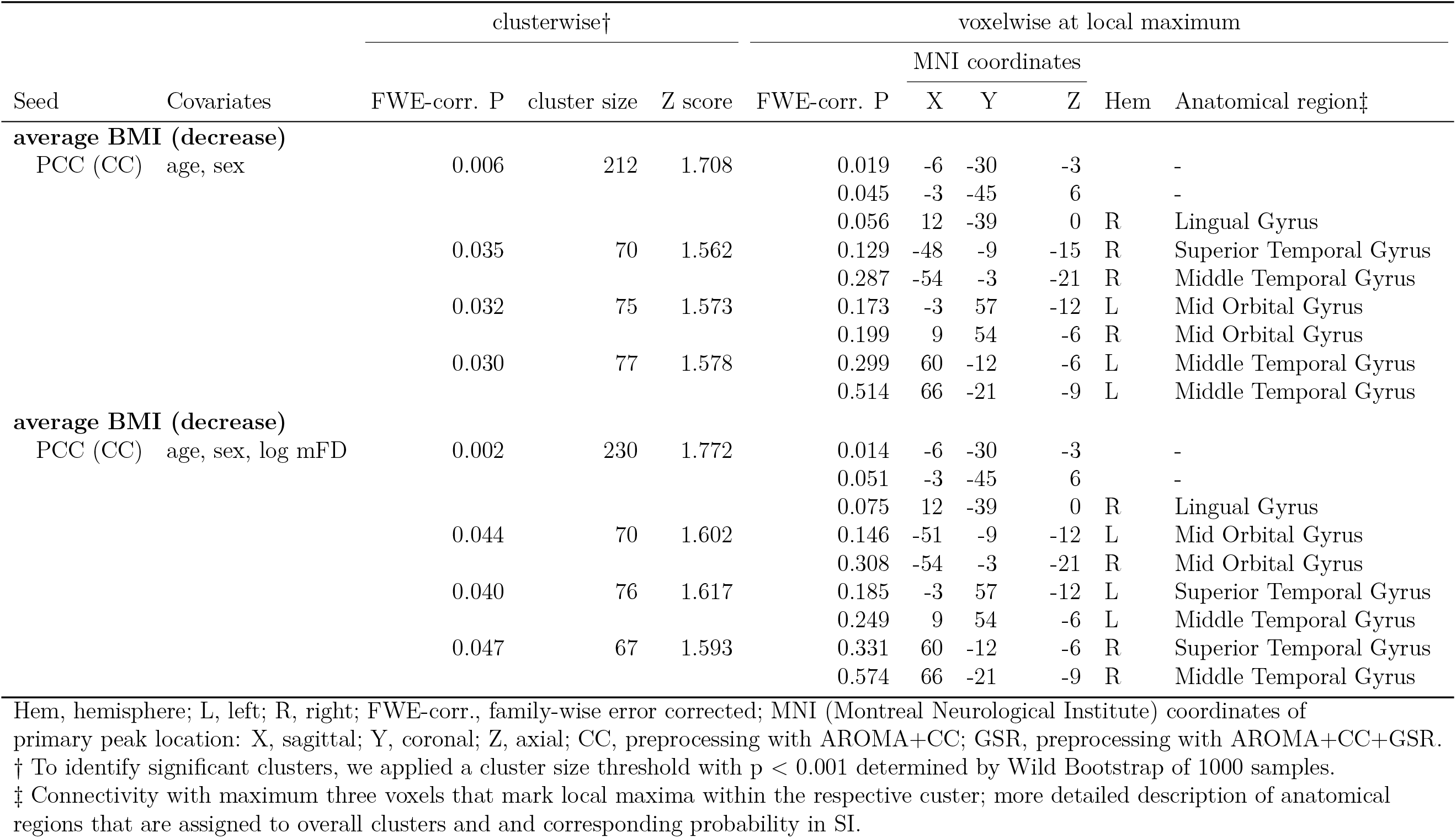
Changes in functional connectivity in whole brain analysis in models EA1 and EA2

**Figure 4.**
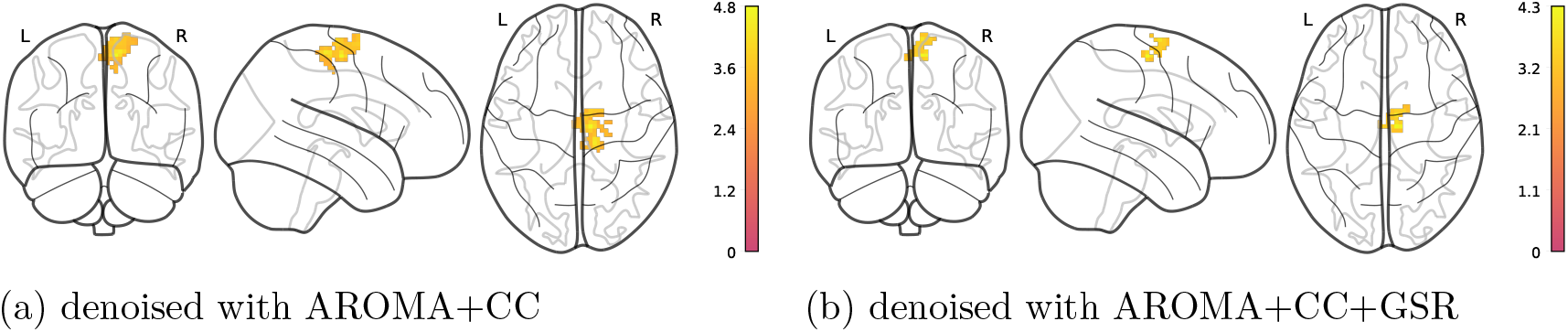
Higher average logmFD is positively associated with FC between NAcc and motor cortex, adjusted for age, sex, average BMI, change in BMI, and change in logmFD (model EA3). Note that clusters have different sizes depending on denoising pipeline. Legends denote empirical Z-values.

**Figure 5.**
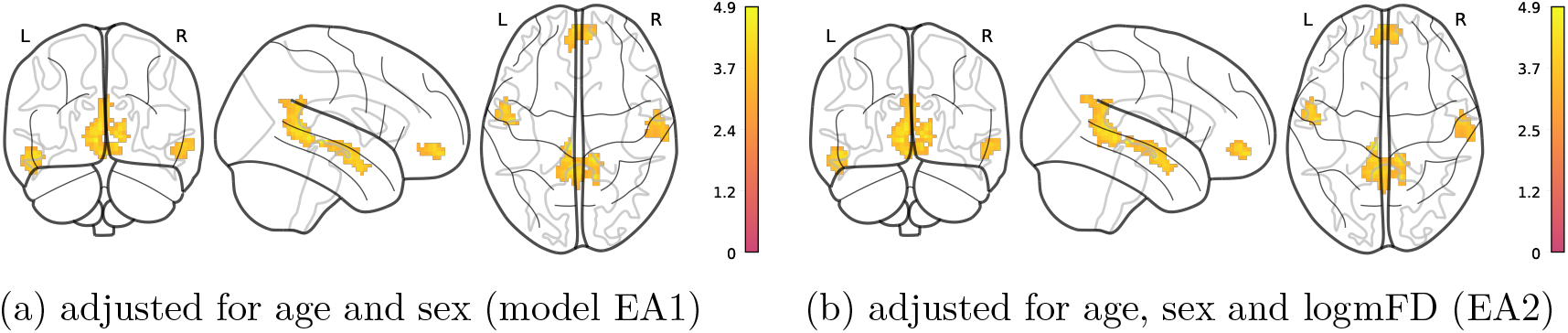
Higher average BMI is associated with lower FC of PCC/precuneus with different regions in AROMA+CC denoised data. Legends denote empirical Z-values.

**Figure 6.**
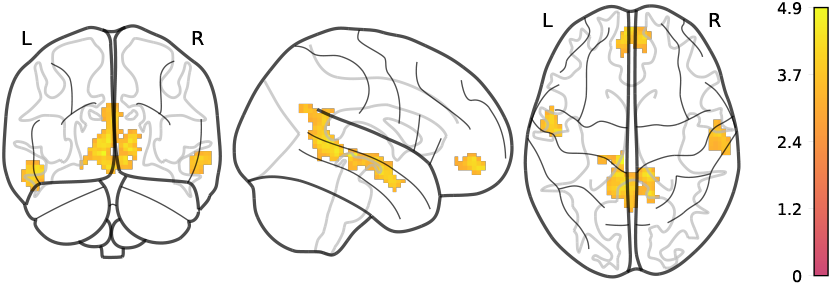
Higher average BMI is associated with lower FC of PCC/precuneus with different regions, adjusted for age, sex, average BMI, average logmFD, and change in logmFD (model EA3) in AROMA+CC denoised data. Legend denotes empirical Z-values.

In the additional exploratory model including only head motion (EA6), higher average logmFD was associated with stronger FC between the NAcc and a cluster located in proximity to the central sulcus and motor areas (see Table 3 and Figure 7). This cluster only differed in size between denoising pipelines. We did not find any clusters with a significant association of either average logmFD or change in logmFD and DMN FC.

**Table 3.** Changes in functional connectivity in whole brain analysis in model EA6

|  |  | clusterwise† |  |  | voxelwise at local maximum |  |  |  |  |  |
| --- | --- | --- | --- | --- | --- | --- | --- | --- | --- | --- |
| Seed | Covariates | FWE-corr. P | cluster size | Z score | FWE-corr. P | MNI coordinates |  |  | Hem | Anatomical region‡ |
|  |  |  |  |  |  | X | Y | Z |  |  |
| **average logmFD (increase)** |  |  |  |  |  |  |  |  |  |  |
| NAcc (CC) | age, sex | 0.005 | 126 | 1.576 | 0.053 | 9 | -15 | 60 | R | Posterior-Medial Frontal |
|  |  |  |  |  | 0.266 | 12 | -24 | 63 | R | Posterior-Medial Frontal |
|  |  |  |  |  | 0.383 | 9 | -18 | 69 | R | Posterior-Medial Frontal |
| **average logmFD (increase)** |  |  |  |  |  |  |  |  |  |  |
| NAcc (GSR) | age, sex | 0.027 | 54 | 1.770 | 0.138 | 9 | -15 | 60 | R | Posterior-Medial Frontal |
|  |  |  |  |  | 0.406 | 9 | -6 | 72 | R | Posterior-Medial Frontal |
|  |  |  |  |  | 0.898 | 0 | -15 | 60 | L | Posterior-Medial Frontal |

Hem, hemisphere; L, left; R, right; FWE-corr., family-wise error corrected; MNI (Montreal Neurological Institute) coordinates of primary peak location: X, sagittal; Y, coronal; Z, axial; CC, preprocessing with AROMA+CC; GSR, preprocessing with AROMA+CC+GSR.
† To identify significant clusters, we applied a cluster size threshold with $p < 0.001$ determined by Wild Bootstrap of 1000 samples.
‡ Connectivity with maximum three voxels that mark local maxima within the respective cluster; more detailed description of anatomical regions that are assigned to overall clusters and corresponding probability in SI.

**Figure 7.**
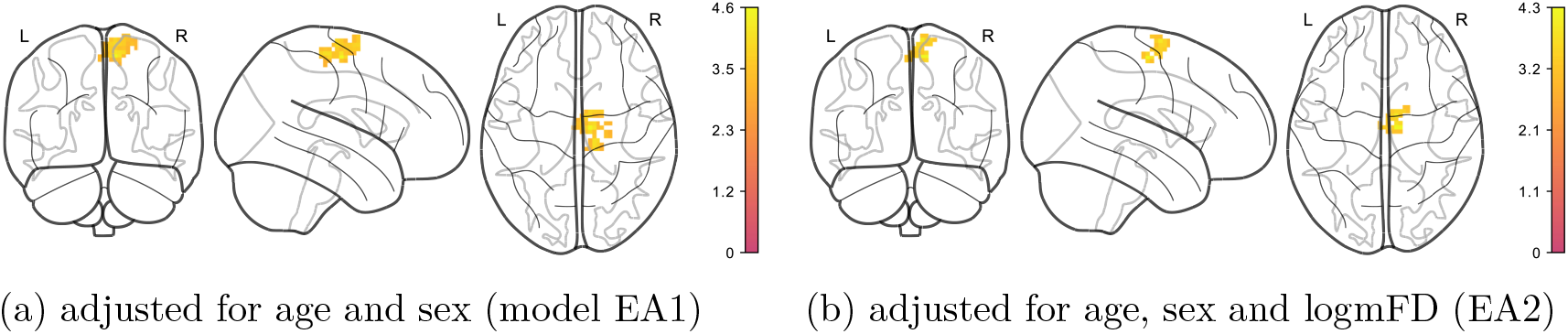
Positive association of average logmFD and FC of NAcc with a cluster in motor cortex, adjusted for age, sex, average BMI, change BMI and change logmFD. Legends denote empirical Z-values.

### Aggregated FC Analysis

As expected, there was no association of average BMI or within-subject BMI change and within RN FC in models EA1, EA2 or EA3 adjusting for age, sex and logmFD (for detailed results see SI).

Like in the whole-brain analysis, higher average BMI was associated with reduced aggregated DMN FC, regardless of whether we adjusted for logmFD (model EA1: *p* = 0.014 and EA2: *p* = 0.017). The association also remained significant when we split logmFD into average and change in logmFD (EA3) (*p* = 0.015) and there was no significant association of average or change in logmFD with DMN FC (see SI for overview of all models).

### Functional Decoding

Functional decoding of the two activation patterns from the pre-registered, exploratory analysis EA3 showed different top association terms. For the https://neurosynth.org/decode/?neurovault=441783 frontal, anterior insula and inferior frontal were the top terms, while for the https://neurosynth.org/decode/?neurovault=441784 primary motor, motor and premotor cortex were the meta-analytic activation maps most similar to this contract. The FC contrasts were thus somewhat distinct, though the decoding method does not allow to conclude specificity (for further information see here https://www.talyarkoni.org/blog/tag/neurosynth/.

## Discussion

In this pre-registered study, we investigated the effects of bariatric surgery on the FC of major resting-state brain networks in a longitudinal controlled design. Moreover, we explored the longitudinal relationship of surgery-induced weight loss and FC, and carefully adjusted for head motion by using two efficient denoising pipelines and controlling for head motion on the group level.

We did not detect significant effects of bariatric surgery compared to waiting on whole-brain FC of the PCC and NAcc, core hubs of the RN and DMN, according to pre-registered whole-brain analyses. This was regardless of whether we adjusted for age, sex and individual head motion. DMN and RN FC was lower at baseline compared to followup for the whole group only when adjusting for age, sex, average logmFD and baseline BMI. In an exploratory model disentangling the effects of average and change in BMI, higher BMI was associated with lower DMN FC for the more lenient denoising pipeline. When we additionally adjusted for both, average and change in head motion, decreases in BMI between the three time points were associated with increased connectivity of the NAcc with a posterior-medial frontal cluster. This result was significant in both denoising pipelines. Functional decoding revealed similarities of the connectivity pattern with frontal, anterior insula and inferior frontal activation patterns. Finally, higher average head motion was associated with increased NAcc connectivity with a cluster in precentral gyrus, close to, yet more posterior cluster associated with change BMI.

In this study, we could not confirm our pre-registered hypotheses. Based on previous studies in bariatric surgery patients, we expected within-DMN FC to increase, and DMN FC to other somatosensory and attention networks to decrease, in line with more efficient processing of visceral and bodily signals after surgery (Frank et al., 2013; Li et al., 2018; McFadden et al., 2013). Further, we expected FC between RN regions to decrease, as bariatric surgery has been previously shown to reduce hyperactivation in RN regions and hedonic motivation to eat (Cerit et al., 2019; Ochner, Stice, et al., 2012; Scholtz et al., 2013). These studies, notably, did not include adequate longitudinal obese control groups, making false-positive findings possible. We thus conclude that surgery-induced heavy weight loss does not strongly affect DMN and RN FC based on the current results.

However, in an exploratory analysis, stronger BMI decrease predicted higher connectivity of the NAcc and a cluster in a posterior-medial frontal brain region. Based on the Harvard-Oxford atlas and NeuroSynth decoding this region might be part of the salience network and involved in action preparation. The enhanced FC between the NAcc and this regions seems at odds with our expectation of reduced hedonic drive to eat after bariatric surgery. On the other hand, higher connectivity might also indicate a better integration of hedonic drive and salience processing in action planning. Previously, a decrease in local (regional homogeneity and frequency of low-amplitude oscillations) and global connectivity (degree centrality) measures in a similar region of the left SMA was reported after glucose administration (Al-Zubaidi et al., 2019). Reduced connectivity in this region was interpreted as an inhibition of action planning or initiation because of fulfilled energy requirements and reduced need for foraging. Importantly, a global decrease in connectivity might go along with selectively increased connectivity, i.e. a shift from integration towards segregation and more specialized processing. Yet, this interpretation is highly speculative, and more studies are needed to corroborate these considerations.

Overall, our results point to the importance of head motion as a confounder in neuroimaging studies in obesity, challenging definite conclusions on the relationship between weight loss and FC changes. Previously, we reported multi-collinearity between BMI and head motion in this sample (Beyer et al., 2020), and therefore conducted careful analyses of the impact of head motion on our results. To our surprise, the effect of weight loss on the connectivity of the NAcc with the posterior-medial frontal region was only present when separating average mFD and change in mFD and thereby introducing two instead of one regressor into the model. These results may be due to the presence of multi-collinearity between change in FD and change in BMI which might appear more pronounced in the split model, and thus lead to unreliable estimations of effects and standard errors. Contrarily, one could argue that only with the careful disentanglement of average and change in BMI and FD the effect of change in BMI could be singled out. This argument is supported by the survival of the cluster when using AROMA+CC+GSR denoised data, and the distinct results of the decoding analysis. Further, average FD was associated with a cluster in a similar, yet not identical region, and crucially, this association was positive. Thus, confounding of the negative BMI change effect and the positive average head motion effect on posterior-frontal FC seems unlikely. Head motion also played a role in the association of higher BMI and reduced DMN FC. While this result was no longer significant on a whole-brain level when using stringent denoising, aggregated within-DMN FC was negatively associated with BMI in both denoising schemes. Thereby, this results echoes a previous finding from our group, and may be interpreted as accelerated age-related decline of the DMN in relation to the cardiometabolic risk related to BMI. Yet, midline regions are prone to motion artifacts and doubts regarding the complete removal of motion confounding remain (Savalia et al., 2016).

The major strength of our study is the prospective intervention controlled design. We compared bariatric surgery patients to an obese control group who did not differ in baseline BMI, comorbidities, treatment history or recommendation and were scanned after the same time intervals (Thiese, 2014). Another strength of our study was the pre-registered analysis plan, which was corrected prior to statistical analysis after we noticed flaws in the first version. In particular, we gave more details on denoising pipelines and models and determined that we would use SwE toolbox, an advanced statistical toolbox to deal with longitudinal repeated measures (Guillaume et al., 2014). Opposed to the flexible factorial models which is the standard in SPM, marginal models use less degrees of freedom, and thus allow for the inclusion of covariates and higher power.

Limitations of our study include the low number of patients who participated in all three time points. In total, only 34 participants contributed to the estimation of the longitudinal effects with at least two time points. Patients in the intervention group were not missing at random over time points, as often, before surgery, they did not fit into the MRI scanner. While this sample size is comparable to previous rsfMRI studies in bariatric surgery, it seems unlikely that our power was high enough to detect small effect sizes. We used seed-based connectivity to derive large-scale brain networks. While this approach yielded reasonable DMN and RN maps, it is a univariate approach not taking into account the inter-relatedness of subnetworks and assuming that the connectivity of a central hub reflects the connectivity of the network as a whole.

Furthermore, our rsfMRI was relatively short, which might have further reduced our power. We did not monitor hunger or satiety in our design, although all participants were scanned after the intake of a breakfast following an overnight fast. Hunger feelings and levels of appetite regulating hormones such as insulin and ghrelin have been shown to predict RN responsivity to food cues, as well as resting-state brain organization (Kroemer et al., 2012; Lepping et al., 2015; Ochner, Laferrére, et al., 2012; Wiemerslage et al., 2016), and might thus have confounded our results (Li et al., 2019). Size and composition of our sample did not allow sex-stratified analyses. However, the disproportionate sex distribution is reflective of the prevalence differences and under-utilization of bariatric surgery by men (Chooi et al., 2019; Fuchs et al., 2015).

## Conclusion

Taken together, this prospective well-controlled study did not confirm previous findings claiming strong effects of bariatric surgery on FC of the RN and DMN in obese patients. Differential changes in head motion adjustment strongly altered rsfMRI neuroimaging results. We thus recommend to rigorously control head motion at acquisition through online monitoring or prospective motion correction or to investigate brain organization with less motion-prone techniques such as task-based fMRI. Pre-registration of concrete and testable hypotheses and publication of null findings as done in the current study would help to increase replicability of the field. Moreover, future studies should include obese control groups, and increase efforts to share and pool valuable patient data into meta-analysis to enhance our understanding of the neural underpinnings of altered gut-brain communication after bariatric surgery.

## Data Availability

The data that support the findings of this study are available on request from the corresponding author, VW. The data are not publicly available due to privacy/ethical restrictions as the information could compromise the privacy of research participants.

https://github.com/fBeyer89/ADI_preproc

https://github.com/hsx1/adi2_rsfMRI

## Data availability

The raw data that support the findings of this study are available on request from the corresponding author, VW. The data are not publicly available due to privacy/ethical restrictions as the information could compromise the privacy of research participants. Unthresholded contrasts maps uploaded on Neurovault are accessible under https://identifiers.org/neurovault.collection:9426. Our source code is publicly available. Preprocessing scripts can be found under https://github.com/fBeyer89/ADI_preproc, and analyses scripts can be found under https://github.com/hsx1/adi2_rsfMRI.

