## Supplementary Information for "Effects of bariatric surgery on functional connectivity of the reward and default mode network: a pre-registered analysis"

### Methods

#### Sample

Table 1 shows the distribution of data points in the present rsfMRI study, for the bariatric surgery group (BARS) and the waiting-list control group (NBARS).

Table 1
*Distribution of data points at months after intervention*

| | BARS | NBARS |
|---|---|---|
| count: only 0 | 7 | 2 |
| count: only 6 | 3 | 0 |
| count: only 12 | 2 | 0 |
| count: 0 and 6 | 4 | 3 |
| count: 6 and 12 | 7 | 1 |
| count: complete data | 10 | 9 |
| total number of subjects | 33 | 15 |
| total data points | 64 | 37 |

#### Denoising Pipelines

We pre-registered two distinct denoising pipelines which were previously shown to efficiently remove motion artifacts in fMRI. In Parkes et al., 2018, ICA-AROMA provided an acceptable trade-off between the mitigation of motion-FC correlation and the introduction of a distance-dependence on motion-FC relationship. Ciric et al., 2017 showed that GSR is very efficient in removing the correlation of head motion and FC, and outperforms ICA-AROMA alone. Yet, it introduces spatial dependency and spurious correlations.

**ICA-AROMA denoising pipeline.** As described in Pruim et al., 2015, the minimally preprocessed data underwent independent component analysis (ICA) using FSL's `melodic` and nuisance components were classified according to four spectral and spatial criteria. Finally, the timeseries of the nuisance components were regressed from the raw rsfMRI timeseries non-aggressively. As recommended in the original paper, we performed regression of WM and CSF signal on the ICA-AROMA result. We used CompCor to estimate 5 variance components from a combined WM and CSF mask, which was further eroded using `fslmaths -nan -thr 0.99 -ero -bin` Behzadi et al., 2007; Muschelli et al., 2014. After regression of these nuisance components from the data, we performed high-pass filtering at 0.01 Hz.

**Global Signal Regression (GSR).** The global signal was derived from the average of all voxels in the brain mask. Then, we regressed WM and CSF CompCor components along with the GS from the ICA-AROMA rsfMRI data and high-pass filtered the resulting file as above.

### Quality Control

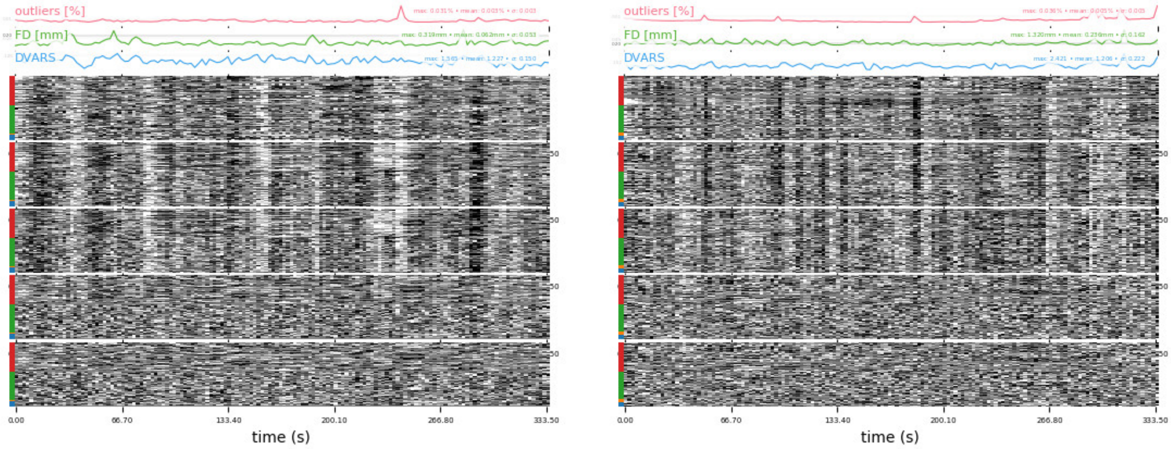


(a) Example 1 with strong amount of structured noise

(b) Example 2 with less amount of structured noise

*Figure 1*. Carpet plot for quality control

**Individual level rsfMRI QC.** On the individual level, we used `plot_carpet` from `niworkflows.viz.plots` to generate carpet plots which depict denoising quality. Figure 1 shows an exemplary carpet plot with (from top to bottom): percent outliers defined by AFNI, mFD, `OutlierCount`, DVARS (spatial root mean square of the data after temporal differencing),voxelwise timeseries from minimally preprocessed, AROMA non-aggressive and aggressive denoising, CC and CC + GSR data (red: GM voxels, green: WM voxels, blue: ventricle, green: cerebellum). All carpet plots were visually checked and if there was still structured noise in the CC+GSR-denoised functional data (such as black/white stripes, signal dropout), the participant was given a rating of 1 in `QA residuals`, and checked again visually. No participant was excluded based on this visual inspection.

**Group level rsfMRI QC.** As described in Beyer et al., 2020, there was a group-by-time interaction on mFD (see Figure 2, upper right panel). The intervention group reduced in head motion in the followup assessments compared to the control group. For this study, we further explored mFD and its relation to DVARS, the root mean square of the temporal change of the voxel-wise signal at each time point. DVARS measures volume-to-volume signal variation. DVARS did not qualitatively differ between groups and time points, and the correlation of DVARS and mFD remained similar over time points (see Figure S2 in suppplements). To further assess the confounding of head motion and FC, we evaluated mFD-FC relationships by computing a functional connectome based on the protocol by Ciric et al., 2018. We used the MNI coordinates of 246 cortical and subcortical spherical seed regions from Power et al., 2012 with 5mm radius in `NiftiSpheresMasker`, and calculated the Spearman rank correlation between these time series. Then, we calculated the distance-dependence of mFD-FC correlations by correlating the euclidean distance between nodes with the mFD-QC correlation.

In the minimally preprocessed data, mFD and FC between nodes were significantly positively correlated (mean Spearman's $r$ = 0.07)(see Table 2). This relationship was negatively associated with distance between nodes (mean Spearman's $r$ = -0.29). Denoising pipelines (AROMA, AROMA+CC, AROMA+CC+GSR) gradually reduced the correlation of mFD and FC, and reduced the distance dependency. Contrary to previous reports, GSR did not exercabate mFD-FC distance dependency Power et al.,

Table 2
*Summary of quality metrics*

| | mean mFD-FC | median mFD-FC | sig. conn. | sig. conn. BH | mean distance-mFDFC | $p_{dist}$ |
|---|---|---|---|---|---|---|
| minimally processed | 0.072 | 0.071 | 7349 | 1571 | -0.292 | < 0.0001 |
| AROMA | 0.061 | 0.063 | 4854 | 20 | -0.021 | 0.0001 |
| AROMA + CC | 0.034 | 0.037 | 5756 | 237 | 0.062 | < 0.0001 |
| AROMA + CC + GSR | 0.006 | 0.010 | 5838 | 407 | 0.069 | < 0.0001 |

mean mFD-FC, mean Spearman's correlation of mFD and FC; median mFD-FC, median Spearman's correlation of mFD and FC; sig. conn., number of significant connections; sig. conn. BH, number of FDR-corrected, significant connections; mean distance-mFDFC, mean Spearman's correlation of distance and mFD-FC correlation; $p_{dist}$, p-value of distance dependency)

2015. Overall, AROMA+CC+GSR performed best, with almost no mFD-FC correlation (mean Spearman's $r = 0.01$), and minimal positive distance dependence (mean Spearman's $r = 0.07$) In addition, for models showing a significant time-by-group interaction, we planned to perform a sensitivity analysis excluding the 10% data sets with highest mFD.

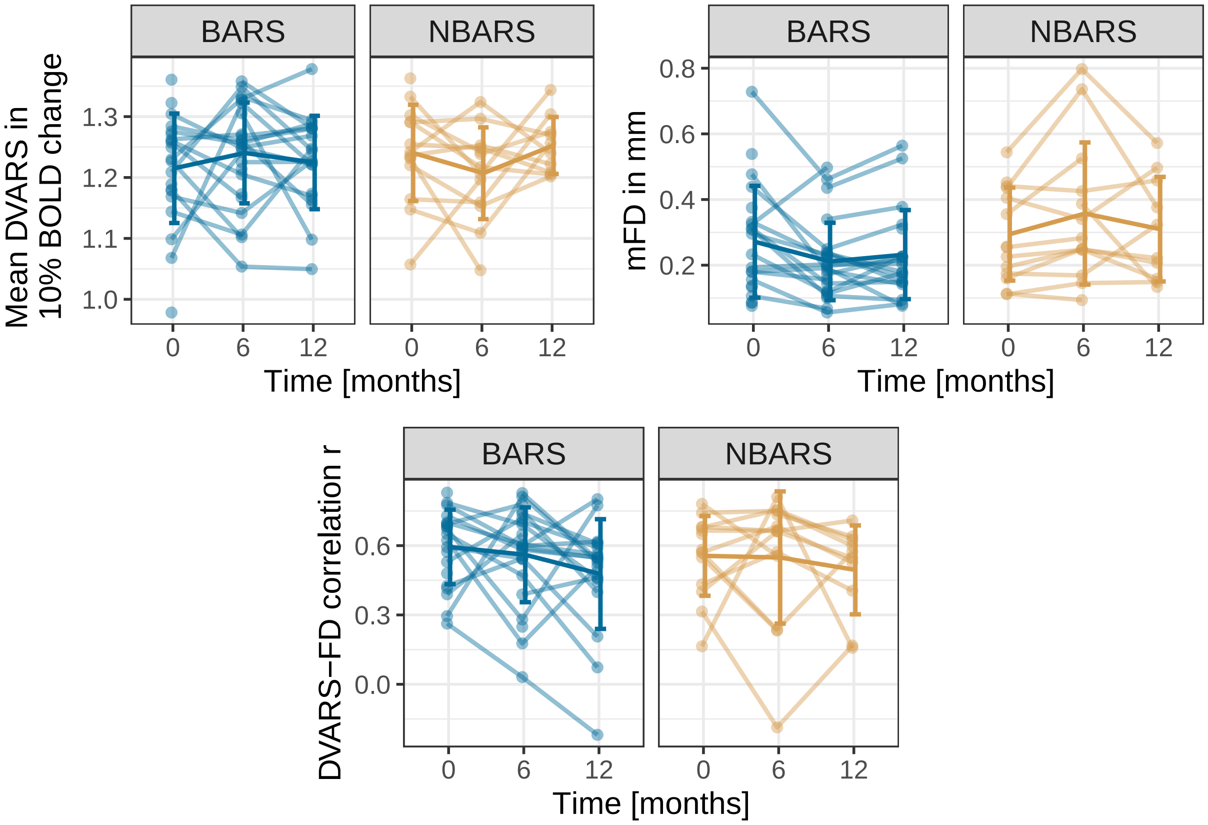


*Figure 2*. Summary of quality measures DVARS, mFD and their correlation

Figure 3 shows the distribution of FC-QC correlations for minimally preprocessed, AROMA, AROMA+CC and AROMA+CC+GSR preprocessed data. With more

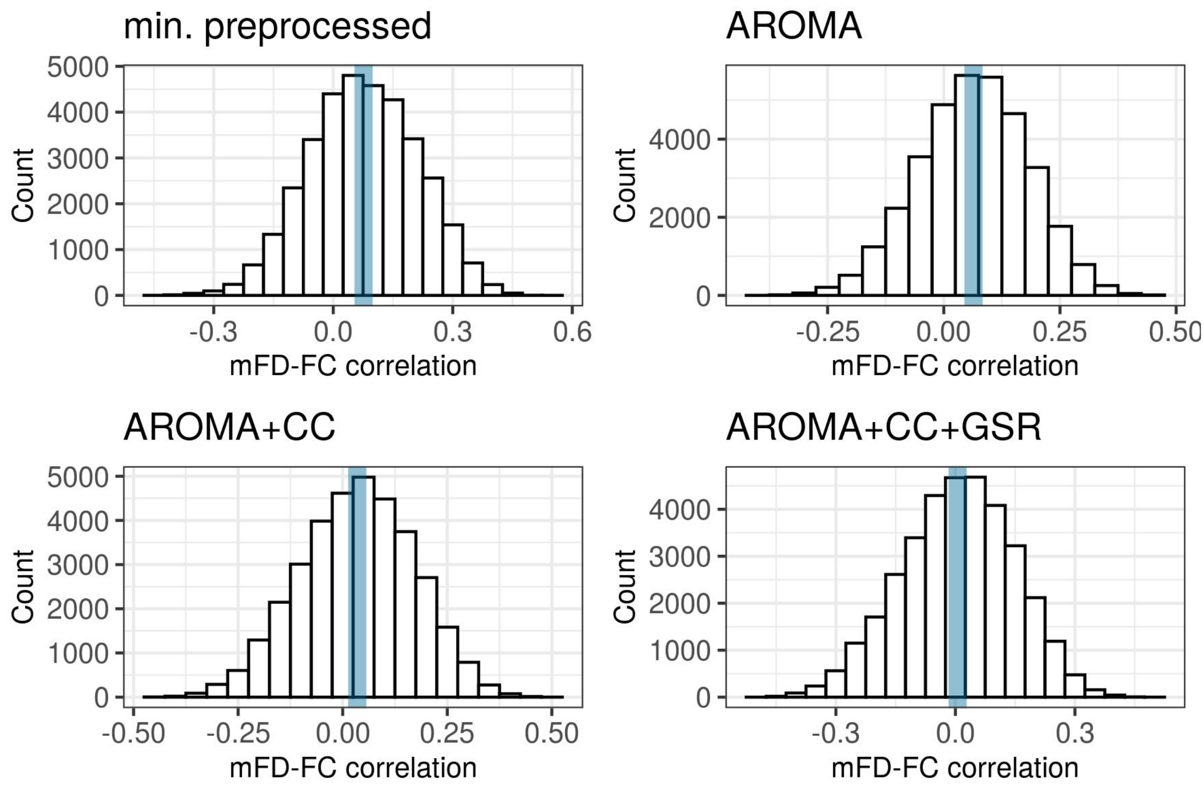


*Figure 3*. Histograms of FC-QC correlations for different denoising pipelines)

denoising, there is a decreasing number of high correlations between FC and the measure of head motion, supporting the positive effect of denoising.

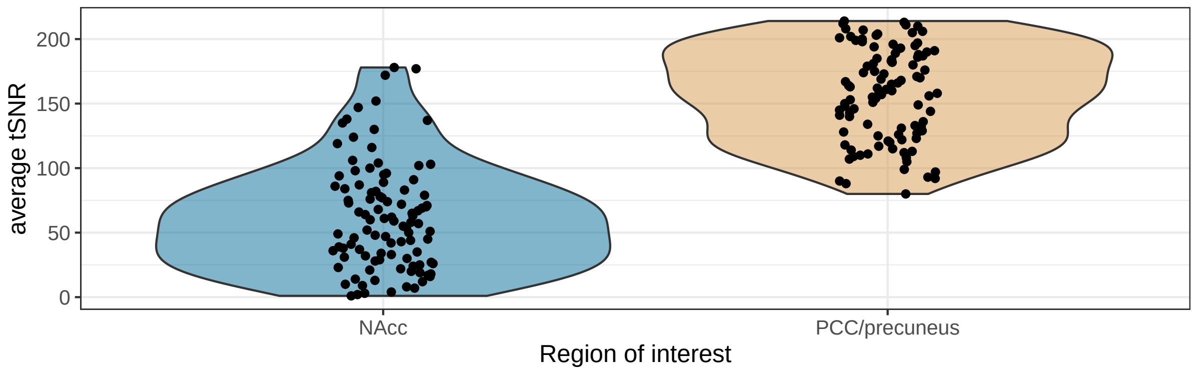


*Figure 4*. Lower tSNR in the region of interest NAcc versus precuneus used for seed-based connectivity (shown are all 101 time points)

### DMN and RN Networks

Figure 5 shows the DMN and reward network average maps which we used for extracting aggregated FC values. Both networks included the typically described brain regions (i.e. medial prefrontal cortex, parietal lobules and posterior hippocampus for the DMN, and anterior cingulate, right amygdala and ventromedial-prefrontal cortex for the reward network). Yet, the reward network was less pronounced (overall lower T-values) and less symmetric (no significant connectivity with left amygdala), which might be due to lower SNR in the seeded brain region (see 4). Unthresholded maps for the t-tests of the DMN and reward network (which were used for data aggregation) are on Neurovault.

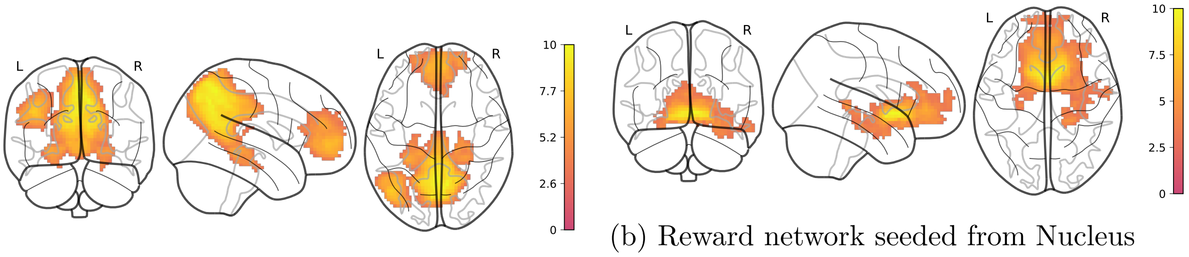


(a) DMN seeded from precuneus

(b) Reward network seeded from Nucleus Accumbens

*Figure 5*. Resting state networks based on AROMA+CC+GSR input data, one-sample t-tests adjusting for age and sex over all time points and participants with bootstrapped clusterwise inference (FWE-corrected $p < 0.05$). Legend denotes empirical Z-values.

### Statistical analysis

For our confirmatory analysis on the model CA3 adjusting age, sex, average logmFD and baseline BMI, one participant in the intervention group had no baseline BMI. In order to make full use of the data available, we employed multivariate imputation, using the package `mice` based on subject identification, condition, baseline age, and available BMI values to obtain a reasonable estimate for the missing baseline BMI. The package allows taking into account the multi-level structure of the data and averaging across multiple possible values, in this case 50 different values, each derived from 10 iterations. Figure 6 illustrates the design matrix used for the confirmatory analyses (regressors for confounders not depicted).

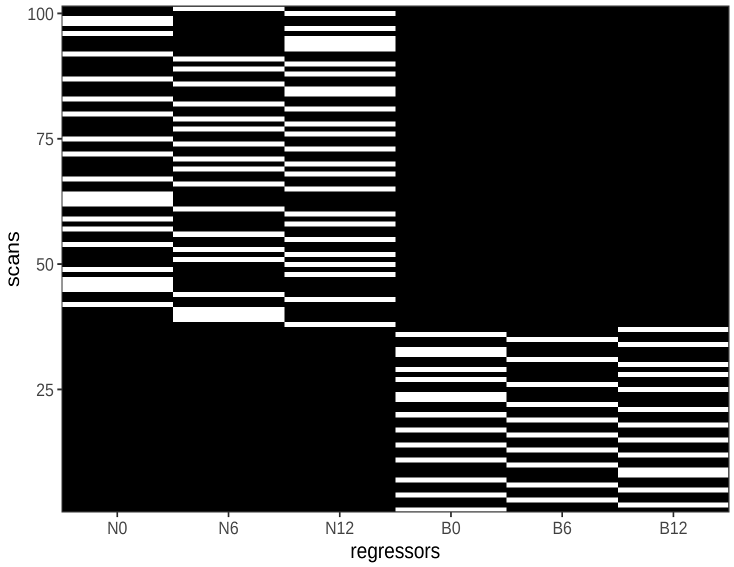


*Figure 6*. Design matrices of time-by-group model with time as categorial factor. N, NBARS; B, BARS; numbers indicate month of measurement since beginning, 0 indicates baseline(pre-surgery measurement, 6 and 12 indicate months after baseline/surgery.

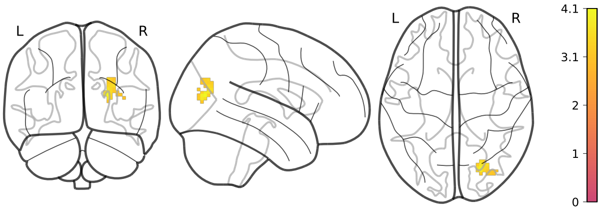


*Figure 7*. Main effect of time: Negative Association of FC between NAcc and this cluster in lateral parietal cortex, adjusted for age, sex, avgFD, and baseline BMI in AROMA+CC+GSR preprocessed data. Legend denotes empirical Z-values.

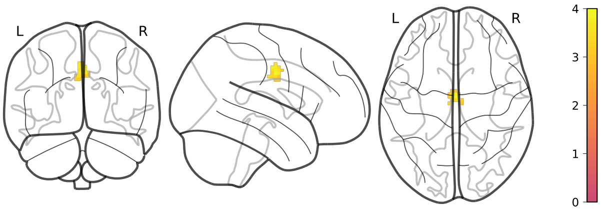


*Figure 8*. Main effect of time: Negative Association of FC between PCC/precuneus and this cluster in ACC, adjusted for age, sex, avgFD, and baseline BMI in AROMA+CC+GSR preprocessed data. Legend denotes empirical Z-values.

### Results

#### Whole brain analysis: group-by-time interaction effect

#### Aggregated FC: group-by-time interaction effect

**Reward network.** Illustration of empirical trajectories and results from mixed-models with aggregated FC in Table 3 to 5 for two time points and Table 6 to 8.

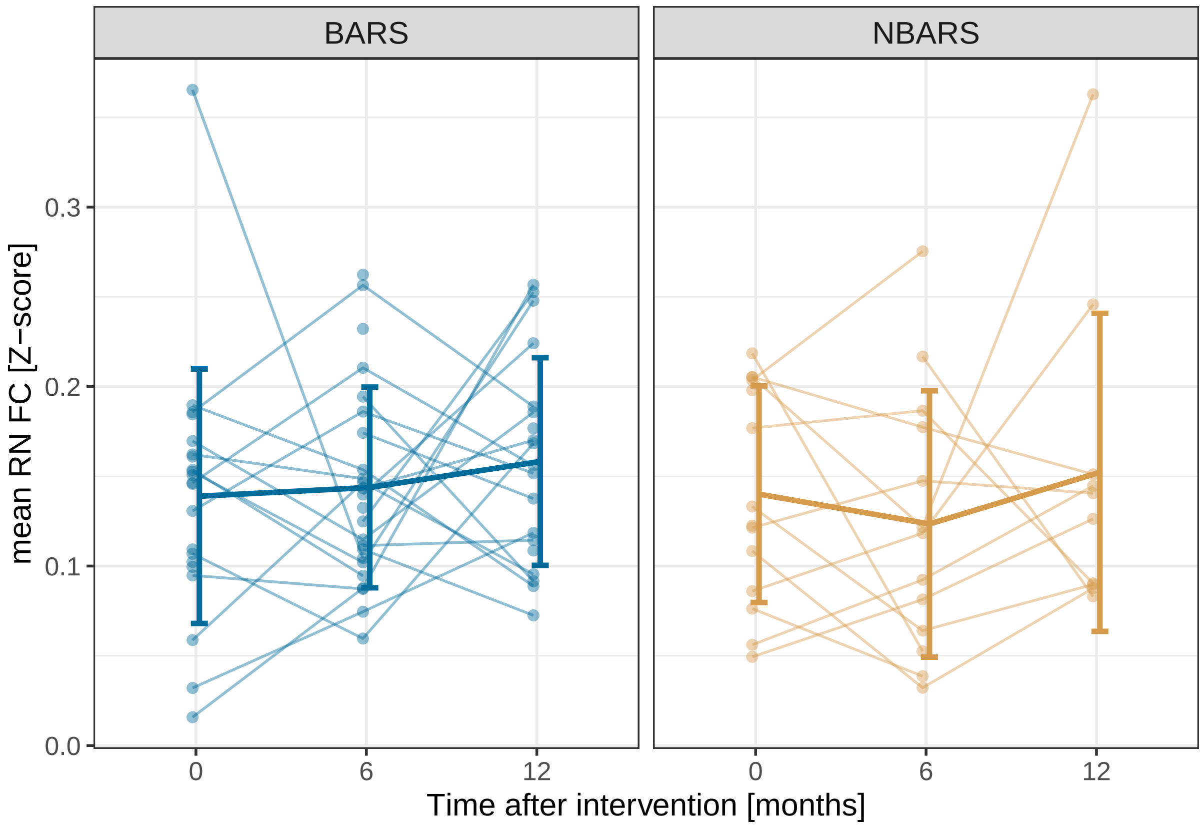


Table 3

*Two time points, without adjustments (model CA1)*

| | R0 | R1 |
|---|---|---|
| Intercept (BL, NBARS) | 0.136 (0.015)*** | 0.142 (0.017)*** |
| Timepoint FU | −0.006 (0.013) | −0.018 (0.021) |
| Group BARS | 0.009 (0.017) | −0.000 (0.022) |
| Timepoint x Group | | 0.019 (0.027) |
| AIC | −165.366 | −158.506 |
| Log Likelihood | 87.683 | 85.253 |
| Num. obs. | 72 | 72 |
| Num. groups: subj.ID | 46 | 46 |
| Var: subj.ID (Intercept) | 0.001 | 0.001 |
| Var: Residual | 0.003 | 0.003 |

Standard deviation is shown in brackets.
$^{***}p < 0.001$; $^{**}p < 0.01$; $^{*}p < 0.05$

**DMN.** Illustration of empirical trajectories and results from mixed-models with aggregated FC in Table 9 to 11 for two time points and Table 12 to 11.

Table 4

*Two time points, adjusted for age, sex and average logmFD (model CA2)*

| | R0 | R1 |
|---|---|---|
| Intercept (BL, NBARS) | 0.082 (0.056) | 0.091 (0.057) |
| Timepoint FU | −0.007 (0.014) | −0.018 (0.021) |
| Group BARS | 0.007 (0.019) | −0.003 (0.024) |
| Timepoint x Group | | 0.019 (0.028) |
| Age | 0.001 (0.001) | 0.001 (0.001) |
| Sex | −0.003 (0.011) | −0.004 (0.011) |
| av logmFD | −0.035 (0.043) | −0.035 (0.043) |
| AIC | −136.181 | −129.321 |
| Log Likelihood | 76.090 | 73.660 |
| Num. obs. | 72 | 72 |
| Num. groups: subj.ID | 46 | 46 |
| Var: subj.ID (Intercept) | 0.001 | 0.001 |
| Var: Residual | 0.003 | 0.003 |

Standard deviation is shown in brackets.

$^{***}p < 0.001$; $^{**}p < 0.01$; $^{*}p < 0.05$

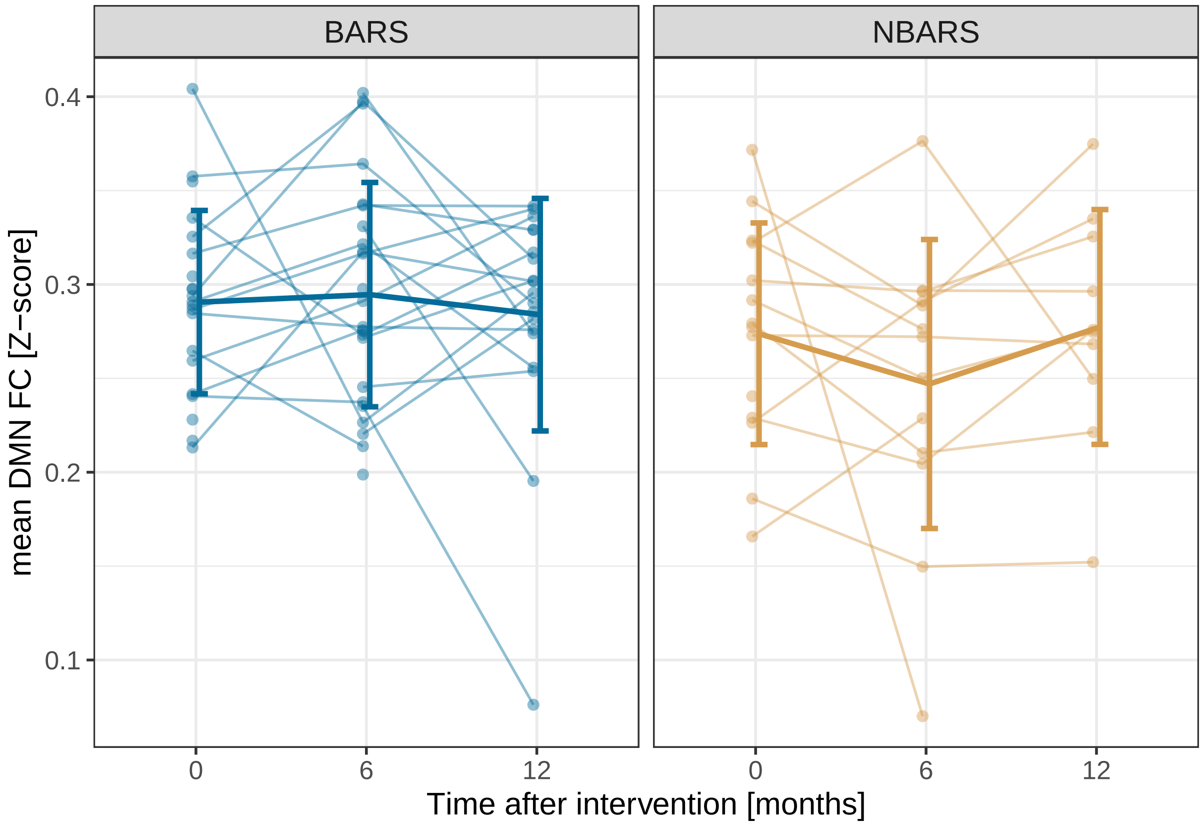

Table 5

*Two time points, adjusted for age, sex, average of logmFD and baseline BMI (model CA3)*

| | R0 | R1 |
|---|---|---|
| Intercept (BL, NBARS) | 0.039 (0.091) | 0.055 (0.094) |
| Timepoint FU | −0.009 (0.014) | −0.018 (0.021) |
| Group BARS | 0.005 (0.019) | −0.003 (0.024) |
| Timepoint x Group | | 0.017 (0.028) |
| Age | 0.001 (0.001) | 0.001 (0.001) |
| Sex | −0.002 (0.011) | −0.003 (0.011) |
| av logmFD | −0.035 (0.044) | −0.034 (0.044) |
| baseline BMI | 0.001 (0.002) | 0.001 (0.002) |
| AIC | −123.570 | −116.611 |
| Log Likelihood | 70.785 | 68.305 |
| Num. obs. | 72 | 72 |
| Num. groups: subj.ID | 46 | 46 |
| Var: subj.ID (Intercept) | 0.001 | 0.001 |
| Var: Residual | 0.003 | 0.003 |

Standard deviation is shown in brackets.
$^{***}p < 0.001$; $^{**}p < 0.01$; $^{*}p < 0.05$

### Exploratory Analyses: Effects of average and change BMI on FC

**Reward network.** Find the results from all mixed-models with aggregated FC in Table 15 and Table 16.

**DMN.** Find the results from all mixed-models with aggregated FC in Table 17 and Table 18.

***Anatomical Labelling for models.*** Find in the detailed Output of the SPM Anatomy toolbox, version 2.2c. For Clusters without any labels, the toolbox was unable to assign any grey matter area. Models evaluated were model EA1 estimating the effects of average BMI and change in BMI, adjusting for age and sex (Table 19), then model EA2 where we additionally adjusted for logmFD (Table 20), and model EA3 estimating the effects of average BMI and BMI variability as well as average logmFD and change in logmFD, adjusting for age and sex (Table 21 to Table 25). Last, the FD model estimating the unique effects of average logmFD and change in logmFD when adjusting for age and sex was evaluated (Table 26 and Table 27).

Table 6
*Three time points, without adjustments (model CA1)*

| | R0 | R1 |
|---|---|---|
| Intercept (BL, NBARS) | 0.135 (0.014)*** | 0.140 (0.018)*** |
| Timepoint FU | −0.004 (0.015) | −0.016 (0.024) |
| Timepoint FU2 | 0.017 (0.016) | 0.013 (0.026) |
| Group BARS | 0.009 (0.015) | −0.001 (0.023) |
| Timepoint FU x Group | | 0.020 (0.031) |
| Timepoint FU2 x Group | | 0.007 (0.033) |
| AIC | −228.437 | −214.503 |
| Log Likelihood | 120.218 | 115.251 |
| Num. obs. | 101 | 101 |
| Num. groups: subj.ID | 48 | 48 |
| Var: subj.ID (Intercept) | 0.001 | 0.001 |
| Var: Residual | 0.004 | 0.004 |

Standard deviation is shown in brackets.
$^{***}p < 0.001$; $^{**}p < 0.01$; $^{*}p < 0.05$

Table 7
*Three time points, adjusted for age, sex and average logmFD (model CA2)*

| | R0 | R1 |
|---|---|---|
| Intercept (BL, NBARS) | 0.089 (0.047) | 0.097 (0.049)* |
| Timepoint FU | −0.004 (0.015) | −0.017 (0.024) |
| Timepoint FU2 | 0.018 (0.016) | 0.013 (0.026) |
| Group BARS | 0.005 (0.016) | −0.006 (0.024) |
| Timepoint FU x Group | | 0.022 (0.031) |
| Timepoint FU2 x Group | | 0.009 (0.033) |
| Age | 0.000 (0.001) | 0.000 (0.001) |
| Sex | −0.005 (0.009) | −0.006 (0.009) |
| av logmFD | −0.037 (0.037) | −0.037 (0.037) |
| AIC | −199.069 | −185.215 |
| Log Likelihood | 108.535 | 103.607 |
| Num. obs. | 101 | 101 |
| Num. groups: subj.ID | 48 | 48 |
| Var: subj.ID (Intercept) | 0.001 | 0.001 |
| Var: Residual | 0.004 | 0.004 |

Standard deviation is shown in brackets.
$^{***}p < 0.001$; $^{**}p < 0.01$; $^{*}p < 0.05$

Table 8
*Three time points, adjusted for age, sex, average of logmFD and baseline BMI (model CA3)*

| | R0 | R1 |
|---|---|---|
| Intercept (BL, NBARS) | 0.087 (0.074) | 0.101 (0.077) |
| Timepoint FU | −0.004 (0.015) | −0.017 (0.024) |
| Timepoint FU2 | 0.018 (0.016) | 0.013 (0.026) |
| Group BARS | 0.004 (0.016) | −0.006 (0.024) |
| Timepoint FU x Group | | 0.022 (0.031) |
| Timepoint FU2 x Group | | 0.009 (0.034) |
| Age | 0.000 (0.001) | 0.000 (0.001) |
| Sex | −0.005 (0.009) | −0.006 (0.009) |
| av logmFD | −0.037 (0.038) | −0.037 (0.038) |
| baseline BMI | 0.000 (0.001) | −0.000 (0.001) |
| AIC | −185.697 | −171.888 |
| Log Likelihood | 102.849 | 97.944 |
| Num. obs. | 101 | 101 |
| Num. groups: subj.ID | 48 | 48 |
| Var: subj.ID (Intercept) | 0.001 | 0.001 |
| Var: Residual | 0.004 | 0.004 |

Standard deviation is shown in brackets.
$^{***}p < 0.001$; $^{**}p < 0.01$; $^{*}p < 0.05$

Table 9
*Two time points, without adjustments (model CA1)*

| | R0 | R1 |
|---|---|---|
| Intercept (BL, NBARS) | 0.265 (0.014)$^{***}$ | 0.274 (0.016)$^{***}$ |
| Timepoint FU | −0.008 (0.014) | −0.027 (0.022) |
| Group BARS | 0.032 (0.015)$^{*}$ | 0.016 (0.021) |
| Timepoint x Group | | 0.032 (0.028) |
| AIC | −172.192 | −166.152 |
| Log Likelihood | 91.096 | 89.076 |
| Num. obs. | 72 | 72 |
| Num. groups: subj.ID | 46 | 46 |
| Var: subj.ID (Intercept) | 0.000 | 0.000 |
| Var: Residual | 0.003 | 0.003 |

Standard deviation is shown in brackets.
$^{***}p < 0.001$; $^{**}p < 0.01$; $^{*}p < 0.05$

Table 10

*Two time points, adjusted for age, sex and average logmFD (model CA2)*

| | R0 | R1 |
|---|---|---|
| Intercept (BL, NBARS) | 0.262 (0.049)*** | 0.277 (0.051)*** |
| Timepoint FU | −0.006 (0.014) | −0.027 (0.022) |
| Group BARS | 0.029 (0.016) | 0.011 (0.022) |
| Timepoint x Group | | 0.036 (0.028) |
| Age | −0.000 (0.001) | −0.000 (0.001) |
| Sex | −0.007 (0.009) | −0.008 (0.010) |
| av logmFD | −0.005 (0.038) | −0.004 (0.038) |
| AIC | −141.808 | −136.102 |
| Log Likelihood | 78.904 | 77.051 |
| Num. obs. | 72 | 72 |
| Num. groups: subj.ID | 46 | 46 |
| Var: subj.ID (Intercept) | 0.000 | 0.001 |
| Var: Residual | 0.003 | 0.003 |

Standard deviation is shown in brackets.
$^{***}p < 0.001$; $^{**}p < 0.01$; $^{*}p < 0.05$

Table 11

*Two time points, adjusted for age, sex, average of logmFD and baseline BMI (model CA3)*

| | R0 | R1 |
|---|---|---|
| Intercept (BL, NBARS) | 0.330 (0.081)*** | 0.367 (0.084)*** |
| Timepoint FU | −0.002 (0.015) | −0.028 (0.022) |
| Group BARS | 0.032 (0.017) | 0.010 (0.022) |
| Timepoint x Group | | 0.043 (0.029) |
| Age | −0.000 (0.001) | −0.000 (0.001) |
| Sex | −0.009 (0.010) | −0.010 (0.010) |
| av logmFD | −0.006 (0.038) | −0.005 (0.038) |
| baseline BMI | −0.002 (0.001) | −0.002 (0.001) |
| AIC | −129.742 | −124.723 |
| Log Likelihood | 73.871 | 72.361 |
| Num. obs. | 72 | 72 |
| Num. groups: subj.ID | 46 | 46 |
| Var: subj.ID (Intercept) | 0.000 | 0.000 |
| Var: Residual | 0.003 | 0.003 |

Standard deviation is shown in brackets.
$^{***}p < 0.001$; $^{**}p < 0.01$; $^{*}p < 0.05$

Table 12
*Three time points, without adjustments (model CA1)*

| | R0 | R1 |
|---|---|---|
| Intercept (BL, NBARS) | 0.268 (0.014)$^{***}$ | 0.275 (0.016)$^{***}$ |
| Timepoint FU | −0.006 (0.013) | −0.028 (0.020) |
| Timepoint FU2 | −0.005 (0.014) | −0.001 (0.022) |
| Group BARS | 0.024 (0.015) | 0.012 (0.021) |
| Timepoint FU x Group | | 0.036 (0.026) |
| Timepoint FU2 x Group | | −0.006 (0.028) |
| AIC | −246.374 | −234.237 |
| Log Likelihood | 129.187 | 125.119 |
| Num. obs. | 101 | 101 |
| Num. groups: subj.ID | 48 | 48 |
| Var: subj.ID (Intercept) | 0.001 | 0.001 |
| Var: Residual | 0.003 | 0.003 |

Standard deviation is shown in brackets.
$^{***}p < 0.001$; $^{**}p < 0.01$; $^{*}p < 0.05$

Table 13
*Three time points, adjusted for age, sex and average logmFD (model CA2)*

| | R0 | R1 |
|---|---|---|
| Intercept (BL, NBARS) | 0.268 (0.046)$^{***}$ | 0.280 (0.048)$^{***}$ |
| Timepoint FU | −0.003 (0.013) | −0.028 (0.020) |
| Timepoint FU2 | −0.003 (0.014) | −0.000 (0.022) |
| Group BARS | 0.020 (0.016) | 0.006 (0.022) |
| Timepoint FU x Group | | 0.040 (0.026) |
| Timepoint FU2 x Group | | −0.003 (0.028) |
| Age | −0.000 (0.001) | −0.000 (0.001) |
| Sex | −0.010 (0.009) | −0.011 (0.009) |
| av logmFD | −0.007 (0.036) | −0.006 (0.037) |
| AIC | −216.912 | −205.185 |
| Log Likelihood | 117.456 | 113.592 |
| Num. obs. | 101 | 101 |
| Num. groups: subj.ID | 48 | 48 |
| Var: subj.ID (Intercept) | 0.001 | 0.001 |
| Var: Residual | 0.003 | 0.003 |

Standard deviation is shown in brackets.
$^{***}p < 0.001$; $^{**}p < 0.01$; $^{*}p < 0.05$

Table 14

*Three time points, adjusted for age, sex, average of logmFD and baseline BMI (model CA3)*

| | R0 | R1 |
|---|---|---|
| Intercept (BL, NBARS) | 0.373 (0.069)*** | 0.399 (0.072)*** |
| Timepoint FU | 0.000 (0.013) | −0.029 (0.020) |
| Timepoint FU2 | 0.001 (0.014) | −0.002 (0.022) |
| Group BARS | 0.027 (0.015) | 0.007 (0.021) |
| Timepoint FU x Group | | 0.048 (0.026) |
| Timepoint FU2 x Group | | 0.008 (0.029) |
| Age | −0.000 (0.001) | −0.000 (0.001) |
| Sex | −0.012 (0.009) | −0.013 (0.009) |
| av logmFD | −0.010 (0.035) | −0.009 (0.035) |
| baseline BMI | −0.003 (0.001)* | −0.003 (0.001)* |
| AIC | −207.304 | −196.179 |
| Log Likelihood | 113.652 | 110.090 |
| Num. obs. | 101 | 101 |
| Num. groups: subj.ID | 48 | 48 |
| Var: subj.ID (Intercept) | 0.001 | 0.001 |
| Var: Residual | 0.003 | 0.003 |

Standard deviation is shown in brackets.
$^{***}p < 0.001$; $^{**}p < 0.01$; $^{*}p < 0.05$

Table 15

*Model EA1 (without adjustment) and EA2 (adjusting for logmFD)*

| | R0 | R1 |
|---|---|---|
| Intercept | 0.164 (0.063)** | 0.118 (0.073) |
| average BMI | −0.001 (0.001) | −0.001 (0.001) |
| change BMI | −0.001 (0.002) | −0.001 (0.002) |
| Age | 0.000 (0.001) | 0.001 (0.001) |
| Sex | −0.009 (0.008) | −0.006 (0.009) |
| logmFD | | −0.040 (0.032) |
| AIC | −203.135 | −197.692 |
| Log Likelihood | 108.568 | 106.846 |
| Num. obs. | 101 | 101 |
| Num. groups: subj.ID | 48 | 48 |
| Var: subj.ID (Intercept) | 0.000 | 0.000 |
| Var: Residual | 0.004 | 0.004 |

Standard deviation is shown in brackets.
$^{***}p < 0.001$; $^{**}p < 0.01$; $^{*}p < 0.05$

Table 16
*Model EA3 (adjusting for average and change in logmFD)*

| | R1 |
|---|---|
| Intercept | 0.123 (0.075) |
| average BMI | −0.001 (0.001) |
| change BMI | −0.000 (0.002) |
| average logmFD | −0.036 (0.036) |
| change logmFD | −0.059 (0.071) |
| Age | 0.000 (0.001) |
| Sex | −0.006 (0.009) |
| AIC | −192.543 |
| Log Likelihood | 105.271 |
| Num. obs. | 101 |
| Num. groups: subj.ID | 48 |
| Var: subj.ID (Intercept) | 0.000 |
| Var: Residual | 0.004 |

Standard deviation is shown in brackets.
$^{***}p < 0.001$; $^{**}p < 0.01$; $^{*}p < 0.05$

Table 17
*Model EA1 (without adjustment) and EA2 (adjusting for logmFD)*

| | R0 | R1 |
|---|---|---|
| Intercept | 0.423 (0.060)$^{***}$ | 0.397 (0.070)$^{***}$ |
| average BMI | −0.003 (0.001)$^{*}$ | −0.003 (0.001)$^{*}$ |
| change BMI | −0.000 (0.001) | 0.000 (0.002) |
| Age | −0.000 (0.001) | −0.000 (0.001) |
| Sex | −0.013 (0.008) | −0.012 (0.008) |
| logmFD | | −0.023 (0.030) |
| AIC | −227.166 | −220.547 |
| Log Likelihood | 120.583 | 118.274 |
| Num. obs. | 101 | 101 |
| Num. groups: subj.ID | 48 | 48 |
| Var: subj.ID (Intercept) | 0.001 | 0.001 |
| Var: Residual | 0.003 | 0.003 |

Standard deviation is shown in brackets.
$^{***}p < 0.001$; $^{**}p < 0.01$; $^{*}p < 0.05$

Table 18

*Model EA3 (adjusting for average and change in logmFD)*

| | R1 |
|---|---|
| Intercept | 0.413 (0.073)*** |
| average BMI | −0.003 (0.001)* |
| change BMI | 0.001 (0.002) |
| average logmFD | −0.009 (0.034) |
| change logmFD | −0.064 (0.059) |
| Age | −0.000 (0.001) |
| Sex | −0.013 (0.008) |
| AIC | −215.664 |
| Log Likelihood | 116.832 |
| Num. obs. | 101 |
| Num. groups: subj.ID | 48 |
| Var: subj.ID (Intercept) | 0.001 |
| Var: Residual | 0.003 |

Standard deviation is shown in brackets.
$^{***}p < 0.001$; $^{**}p < 0.01$; $^{*}p < 0.05$

Table 19

*Anatomical labels for clusters functionally connected to the PCC showing a negative association with average BMI in model EA1 on AROMA+CC denoised data.*

| Cluster | Number of voxels in cluster | % of cluster volume assigned | Hemisphere | Area | % of area overlap with cluster |
|---|---|---|---|---|---|
| Cluster 1 (212 voxel) | | | | | |
| | 5.5 | 2.6 | left | Lobule I IV (Hem) | 3.3 |
| | 2.6 | 1.2 | right | Area hOc1 [V1] | 0.4 |
| | 1.4 | 0.7 | left | Lobule V (Hem) | 0.6 |
| | 0.7 | 0.3 | right | Area hOc2 [V2] | 0.2 |
| | 0.4 | 0.2 | left | Thal: Prefrontal | 0.2 |
| | 0.4 | 0.2 | left | Subiculum | 0.3 |
| | 0.2 | 0.1 | right | Subiculum | 0.2 |
| | 0.2 | 0.1 | left | Thal: Parietal | 0.2 |
| | 0.1 | 0.1 | left | Area hOc1 [V1] | 0.0 |
| | 11.5 | 5.4 | | assigned in total | |
| Cluster 2 (77 voxel) | | | | | |
| | 21.8 | 28.3 | right | Area TE 3 | 7.1 |
| | 21.8 | 28.3 | | assigned in total | |
| Cluster 3 (75 voxel) | | | | | |
| | 37.1 | 49.5 | left | Area Fp2 | 17.3 |
| | 25.9 | 34.5 | right | Area Fp2 | 14.3 |
| | 1.6 | 2.2 | left | Area s32 | 2.6 |
| | 0.6 | 0.7 | right | Area Fp1 | 0.1 |
| | 65.1 | 86.9 | | assigned in total | |
| Cluster 4 (70 voxel) | | | | | |
| | 4.1 | 5.9 | left | Area TE 3 | 1.6 |
| | 0.6 | 0.8 | left | Area Id1 | 1.6 |
| | 4.7 | 6.7 | | assigned in total | |

Table 20
*Anatomical labels for clusters functionally connected to the PCC showing a negative association with average BMI in model EA2 on AROMA+CC denoised data.*

| Cluster | Number of voxels in cluster | % of cluster volume assigned | Hemisphere | Area | % of area overlap with cluster |
|---|---|---|---|---|---|
| Cluster 1 (230 voxel) | | | | | |
| | 5.5 | 2.4 | left | Lobule I IV (Hem) | 3.3 |
| | 2.6 | 1.1 | right | Area hOc1 [V1] | 0.4 |
| | 1.6 | 0.7 | left | Lobule V (Hem) | 0.6 |
| | 1.6 | 0.7 | left | Thal: Prefrontal | 0.7 |
| | 0.7 | 0.3 | left | Thal: Visual | 2.4 |
| | 0.7 | 0.3 | right | Area hOc2 [V2] | 0.2 |
| | 0.5 | 0.2 | left | Thal: Parietal | 0.5 |
| | 0.4 | 0.2 | left | Subiculum | 0.3 |
| | 0.2 | 0.1 | right | Subiculum | 0.2 |
| | 0.1 | 0.1 | left | Area hOc1 [V1] | 0.0 |
| | 13.9 | 6.0 | | assigned in total | |
| Cluster 2 (76 voxel) | | | | | |
| | 37.8 | 49.8 | left | Area Fp2 | 17.6 |
| | 28.4 | 37.4 | right | Area Fp2 | 15.7 |
| | 0.1 | 0.1 | left | Area s32 | 0.1 |
| | 66.3 | 87.2 | | assigned in total | |
| Cluster 3 (70 voxel) | | | | | |
| | 4.1 | 5.9 | left | Area TE 3 | 1.6 |
| | 0.3 | 0.4 | left | Area Id1 | 0.9 |
| | 4.4 | 6.3 | | assigned in total | |
| Cluster 4 (67 voxel) | | | | | |
| | 18.1 | 27.0 | right | Area TE 3 | 5.9 |
| | 18.1 | 27.0 | | assigned in total | |

Table 21
*Anatomical labels for clusters functionally connected to the NAcc showing a negative association with change in BMI in model EA3 on AROMA+CC denoised data.*

| Cluster | Number of voxels in cluster | % of cluster volume assigned | Hemisphere | Area | % of area overlap with cluster |
|---|---|---|---|---|---|
| Cluster 1 (112 voxel) | | | | | |

Table 22
*Anatomical labels for clusters functionally connected to the NAcc showing a positive association with average mFD in model EA3 on AROMA+CC denoised data.*

| Cluster | Number of voxels in cluster | % of cluster volume assigned | Hemisphere | Area | % of area overlap with cluster |
|---|---|---|---|---|---|
| Cluster 1 (143 voxel) | | | | | |
| | 12.9 | 9.0 | right | Area 4a | 4 |
| | 0.1 | 0.1 | left | Area 4a | 0 |
| | 13.0 | 9.1 | | assigned in total | |

Table 23
*Anatomical labels for clusters functionally connected to the NAcc showing a negative association with change in BMI in model EA3 on AROMA+CC+GSR denoised data.*

| Cluster | Number of voxels in cluster | % of cluster volume assigned | Hemisphere | Area | % of area overlap with cluster |
|---|---|---|---|---|---|
| Cluster 1 (99 voxel) | | | | | |

Table 24
*Anatomical labels for clusters functionally connected to the NAcc showing a positive association with average mFD in model EA3 on AROMA+CC+GSR denoised data.*

| Cluster | Number of voxels in cluster | % of cluster volume assigned | Hemisphere | Area | % of area overlap with cluster |
|---|---|---|---|---|---|
| Cluster 1 (99 voxel) | | | | | |

Table 25
*Anatomical labels for clusters functionally connected to the PCC showing a negative association with average BMI in model EA3 on AROMA+CC denoised data.*

| Cluster | Number of voxels in cluster | % of cluster volume assigned | Hemisphere | Area | % of area overlap with cluster |
|---|---|---|---|---|---|
| Cluster 1 (251 voxel) | | | | | |
| | 6.0 | 2.4 | left | Lobule I IV (Hem) | 3.7 |
| | 4.6 | 1.8 | left | Thal: Parietal | 4.3 |
| | 2.7 | 1.1 | right | Area hOc1 [V1] | 0.4 |
| | 2.2 | 0.9 | left | Lobule V (Hem) | 0.9 |
| | 1.8 | 0.7 | left | Subiculum | 1.4 |
| | 1.6 | 0.6 | left | Thal: Prefrontal | 0.7 |
| | 0.7 | 0.3 | right | Area hOc2 [V2] | 0.2 |
| | 0.7 | 0.3 | left | Thal: Visual | 2.4 |
| | 0.2 | 0.1 | right | Subiculum | 0.2 |
| | 0.1 | 0.1 | left | Area hOc1 [V1] | 0.0 |
| | 20.5 | 8.2 | | assigned in total | |
| Cluster 2 (91 voxel) | | | | | |
| | 42.0 | 46.2 | left | Area Fp2 | 19.6 |
| | 32.9 | 36.1 | right | Area Fp2 | 18.1 |
| | 1.6 | 1.8 | left | Area s32 | 2.6 |
| | 76.5 | 84.1 | | assigned in total | |
| Cluster 3 (70 voxel) | | | | | |
| | 4.1 | 5.9 | left | Area TE 3 | 1.6 |
| | 0.3 | 0.4 | left | Area Id1 | 0.9 |
| | 4.4 | 6.3 | | assigned in total | |
| Cluster 4 (69 voxel) | | | | | |
| | 19.8 | 28.7 | right | Area TE 3 | 6.4 |
| | 19.8 | 28.7 | | assigned in total | |

Table 26
*Anatomical labels for clusters functionally connected to the NAcc showing a positive association with average mFD in model EA6 on AROMA+CC denoised data.*

| Cluster | Number of voxels in cluster | % of cluster volume assigned | Hemisphere | Area | % of area overlap with cluster |
|---|---|---|---|---|---|
| Cluster 1 (126 voxel) | | | | | |
| | 5.6 | 4.5 | right | Area 4a | 1.7 |
| | 5.6 | 4.5 | | assigned in total | |

Table 27
*Anatomical labels for clusters functionally connected to the NAcc showing a positive association with average mFD in model EA6 on AROMA+CC denoised data.*

| Cluster | Number of voxels in cluster | % of cluster volume assigned | Hemisphere | Area | % of area overlap with cluster |
|---|---|---|---|---|---|
| Cluster 1 (126 voxel) | | | | | |
| | 5.6 | 4.5 | right | Area 4a | 1.7 |
| | 5.6 | 4.5 | | assigned in total | |
